# Novel CSF tau biomarkers can be used for disease staging of sporadic Alzheimer’s disease

**DOI:** 10.1101/2023.07.14.23292650

**Authors:** Gemma Salvadó, Kanta Horie, Nicolas R. Barthélemy, Jacob W. Vogel, Alexa Pichet Binette, Charles D. Chen, Andrew J Aschenbrenner, Brian A. Gordon, Tammie L.S. Benzinger, David M. Holtzman, John C. Morris, Sebastian Palmqvist, Erik Stomrud, Shorena Janelidze, Rik Ossenkoppele, Suzanne E. Schindler, Randall J. Bateman, Oskar Hansson

## Abstract

Biological staging of individuals with Alzheimer’s disease (AD) may improve diagnostic and prognostic work-up of dementia in clinical practice and the design of clinical trials. Here, we created a staging model using the Subtype and Stage Inference (SuStaIn) algorithm by evaluating cerebrospinal fluid (CSF) amyloid-β (Aβ) and tau biomarkers in 426 participants from BioFINDER-2, that represent the entire spectrum of AD. The model composition and main analyses were replicated in 222 participants from the Knight ADRC cohort. SuStaIn revealed in the two cohorts that the data was best explained by a single biomarker sequence (one subtype), and that five CSF biomarkers (ordered: Aβ42/40, tau phosphorylation occupancies at the residues 217 and 205 [pT217/T217 and pT205/T205], microtubule-binding region of tau containing the residue 243 [MTBR-tau243], and total tau) were sufficient to create an accurate disease staging model. Increasing CSF stages (0-5) were associated with increased abnormality in other AD-related biomarkers, such as Aβ- and tau-PET, and aligned with different phases of longitudinal biomarker changes consistent with current models of AD progression. Higher CSF stages at baseline were associated with higher hazard ratio of clinical decline. Our findings indicate that a common pathophysiologic molecular pathway develops across all AD patients, and that a single CSF collection is sufficient to reliably indicate the presence of both AD pathologies and the degree and stage of disease progression.

## Introduction

Currently, more than 50 million people are affected by dementia and this number is expected to more than double by 2050^1^. Alzheimer’s disease (AD) is the most common form of dementia, characterized by the accumulation of extracellular plaques containing amyloid-β (Aβ) and intracellular tau aggregates in the forms of tau tangles and neuropil threads^2^. Over the last two decades, the AD field has moved towards the use of biomarkers to support the diagnostic and prognostic work-up, rather than relying solely on clinical symptoms^3^. This has been made possible by advancements of imaging and fluid biomarkers that accurately track AD pathology *in vivo*. Given that the accumulation of pathology can take many years to decades^3^, before any clinical symptoms appear, the use of biomarkers is critical to ensuring an early and reliable detection of AD^4^. Key biomarkers may help to improve patient diagnosis, management and prognosis^5–8^. And the use of AD biomarkers will be even more important when disease-modifying treatments become widely available ^9–11^. In this context, a more sophisticated personalized medicine approach of AD, based on high performing AD biomarkers, will become crucial to select the most optimal participants for specific treatments and for enrolment in new clinical trials.

In recent years, multiple cerebrospinal fluid (CSF) biomarkers targeting different pathophysiological mechanisms have been developed (see ^4^ for a review). There has been an increasing interest in developing biomarkers for measuring tau species phosphorylated at different residues. Among the phosphorylated tau (p-tau) species, p-tau181^12–17^, p-tau217^12,13,15,18,19^, and p-tau231^15,20–22^ or the phosphorylation occupancies have been studied in depth, and have shown strong associations with Aβ pathology and moderate associations with tau (as measured by both PET^18,23^ and neuropathology^24,25^). These biomarkers have shown their utility on improving the diagnostic work-up of AD and the prediction of disease progression^12,13,19,26,27^. Other biomarkers, such as p-tau205 or the occupancy (pT205/T205)^28–30^ and microtubule binding region (MTBR) of tau containing the 243 residue (MTBR-tau243)^31,32^, have been more closely related to tau tangle pathology. Importantly, some of these CSF biomarkers were shown to become abnormal at different phases during the progression of autosomal dominant AD (ADAD)^29^, suggesting a sequence of CSF biomarker changes that may serve as a measurable biological indicator tracking advancing disease progression.

The progression of Aβ- or tau-pathology across the brain has been previously used to stage participants across the AD *continuum*^33–38^. However, these models need at least one Aβ- or tau-PET scan, which is expensive, and requires specialized personnel and facilities. Further, information of only one pathological measure (*e.g.*, Aβ or tau) can be obtained from these images. On the other hand, CSF biomarkers are less expensive, more accessible, and multiple pathological measures may be obtained from a single sample. Given this, and with the idea that different CSF biomarkers may become abnormal at different stages of the disease, we aimed to generate a data-driven staging scheme for sporadic AD using key CSF tau biomarkers in combination with CSF Aβ42/40. An unresolved question is whether there is a single molecular pathway throughout the AD *continuum*, or whether there are subtypes of AD following different fluid biomarker trajectories, as has been shown for regional spread of insoluble tau tangles^36,39,40^.

To test this, we used Subtype and Stage Inference (SuStaIn)^41^ to model the most likely sequence of CSF biomarker abnormalities that occur along the AD time course. This machine-learning method uses cross-sectional data to order biomarker abnormalities in a probabilistic manner and at the same time addresses possible diverging trajectories of this ordering. With this approach, we staged 426 participants of the Swedish BioFINDER-2 study, ranging from cognitively unimpaired (CU) participants to patients with mild cognitive impairment (MCI) or dementia. This model was used to assign each participant to a fluid biomarker stage, which was subsequently correlated with established AD key features such as Aβ- and tau-PET, cortical thickness, and cognition. Next, we investigated the accuracy of our staging model to predict amyloid (A) and tau (T) status as defined by PET^42^, as well as its potential as a diagnostic tool for distinguishing diagnostic groups. Using longitudinal data, we further determined trajectories of several key AD-biomarkers based on participants baseline fluid biomarker disease stage. We also tested the clinical utility of our novel staging system for predicting clinical progression. Finally, we performed the main analyses in the independent cohort (the Charles F. and Joanne Knight Alzheimer Disease Research Center [Knight-ADRC]), which included 222 participants. Altogether our results suggest that participants in the AD *continuum* progress along a single path and can be biologically staged using a single sample of CSF. This may have important implications in the clinical practice and in the selection of participants for future clinical trials.

## Results

A total of 426 participants of the Swedish BioFINDER-2 study (NCT03174938)^19^ with complete CSF data were included in this study. From these, 80 were cognitively unimpaired Aβ negative (CU−) and 79 cognitively unimpaired Aβ positive (CU+) participants, 88 were diagnosed with MCI and Aβ positive, 100 were diagnosed with AD dementia and Aβ positive (ADD+), and 79 were assessed as non-AD patients (22 were Aβ positive). Demographic information is presented in Table 1. Of these, 220 participants had longitudinal CSF data available (Supplementary Table 1).

**Table 1:** BioFINDER-2 participants’ characteristics. Data is shown as mean(SD) unless otherwise stated. Participants are divided by clinical diagnosis and amyloid status based on their CSF Aβ42/40 levels (Aβ+: <0.080). Only participants who progressed to MCI or dementia patients due to AD etiology were considered to progress. ^a^, 1 participant missing; ^b^, 4 participants missing; ^c^, 175 participants missing; ^d^, 9 participants missing; ^e^, 6 participants missing; ^f^, 4 participants missing; ^g^, 36 participants missing. Abbreviations: Aβ, amyloid-β; AD, Alzheimer’s disease; ADD+, Alzheimer’s disease dementia amyloid positive; CU−, cognitively unimpaired amyloid negative; CU+, cognitively unimpaired amyloid positive; CSF, cerebrospinal fluid; MCI, mild cognitive impairment amyloid positive; nonAD, non-Alzheimer’s related disease; PET, positron emission tomography; ROI, region of interest; SD, standard deviation; SUVR, standardized uptake value ratio.

|  | **All**<br>**(n=426)** | **CU-**<br>**(n=80)** | **CU+**<br>**(n=79)** | **MCI+**<br>**(n=88)** | **ADD+**<br>**(n=100)** | **nonAD**<br>**(n=79)** |
| --- | --- | --- | --- | --- | --- | --- |
| **Age, years** | 71.5 (8.5) | 70.7 (9.5) | 71.1 (9.5) | 72.0 (7.4) | 73.0 (6.9) | 70.2 (9.0) |
| **Women, n(%)** | 211<br>(49.5%) | 39<br>(48.8%) | 40<br>(50.6%) | 38<br>(43.2%) | 56<br>(56.0%) | 38<br>(48.1%) |
| **APOE-ε4**<br>**carriershp,**<br>**n(%)<sup>a</sup>** | 246<br>(57.7%) | 26<br>(32.5%) | 58<br>(73.4%) | 62<br>(70.5%) | 74<br>(74.0%) | 26<br>(32.9%) |
| **Years of**<br>**education<sup>b</sup>** | 12.3 (3.8) | 12.0 (3.2) | 12.2 (3.4) | 12.7 (4.5) | 12.0 (4.0) | 12.7 (3.5) |
| **Amyloid-PET,**<br>**Centiloids<sup>c</sup>** | 37.3<br>(44.2) | -4.48<br>(9.50) | 41.8<br>(36.0) | 69.6<br>(36.0) | 115 (23.3) | 5.67<br>(23.3) |
| **Tau-PET,**<br>**SUVR<sup>d</sup>** | 1.53<br>(0.61) | 1.16<br>(0.08) | 1.23<br>(0.21) | 1.50<br>(0.45) | 2.38<br>(0.58) | 1.16<br>(0.10) |
| **Cortical**<br>**thickness,**<br>**mm<sup>e</sup>** | 2.46<br>(0.16) | 2.56<br>(0.09) | 2.55<br>(0.12) | 2.46<br>(0.13) | 2.32<br>(0.15) | 2.46<br>(0.18) |
| **CSF NfL<sup>f</sup>** | 245 (175) | 147 (79.1) | 189 (153) | 223 (128) | 316 (180) | 333 (222) |
| **mPACC<sup>g</sup>** | -1.62<br>(2.03) | 0.06<br>(0.76) | -0.26<br>(0.78) | -1.88<br>(0.72) | -4.34<br>(1.72) | -1.69<br>(2.03) |
| **Progressed**<br>**to MCI** | 11<br>(2.6%) | 0<br>(0%) | 11<br>(13.9%) | - | - | - |
| **Progressed**<br>**to ADD+** | 41<br>(9.6%) | 0<br>(0%) | 3<br>(3.8%) | 38<br>(43.2%) | - | - |

### CSF staging model

We initially applied SuStaIn to the BioFINDER-2 cohort using the following CSF biomarkers: the Aβ42/40 ratio, the phosphorylated to non-phosphorylated tau ratio of pT205/T205, pT181/T181, pT217/T217, and pT231/T231, as well as the concentrations of MTBR-tau243 and total tau [the residue 151-153]) based on availability and previous literature. Through a process of model optimization (Ext. Data Fig. 1, see Methods for further details), we arrived on a model that excluded pT181/T181 and pT231/T231 due to information redundancy. SuStaIn revealed that a single biomarker sequence best described the progressive abnormality of the selected biomarkers (Ext Data Fig. 1C). The final ordering of the model was the Aβ42/40 ratio, pT217/T217, pT205/T205, MTBR-tau243 and total-tau (Fig. 1A), resulting in a five-stage model (plus stage 0 as a negative biomarker stage). All BioFINDER-2 participants were then classified into one of these biomarker-based disease stages based on their CSF levels, with 124 (29.1%) being at CSF stage 0, 35 (8.2%) at CSF stage 1, 53 (12.4%) at CSF stage 2, 49 (11.5%) at CSF stage 3, 87 (20.4%) at CSF stage 4 and 78 (18.3%) at CSF stage 5. Demographic, genetic, and diagnostic characteristics of these participants is shown in Ext Data Fig. 2. In brief, the CSF biomarker-based was not associated with sex (χ^2^(5)=7.7, p=0.180) or years of education (χ^2^(5)=4.7, p=0.452), but higher CSF stage was associated with older age (χ^2^(5)=16.9, p=0.005), carriership of an *APOE-ε4* allele (χ^2^(5)=72.8, p<0.001) and a more advanced clinical disease stage (χ^2^(5)=478.6, p<0.001, Ext Data Fig. 2A-E).

**Fig 1:**
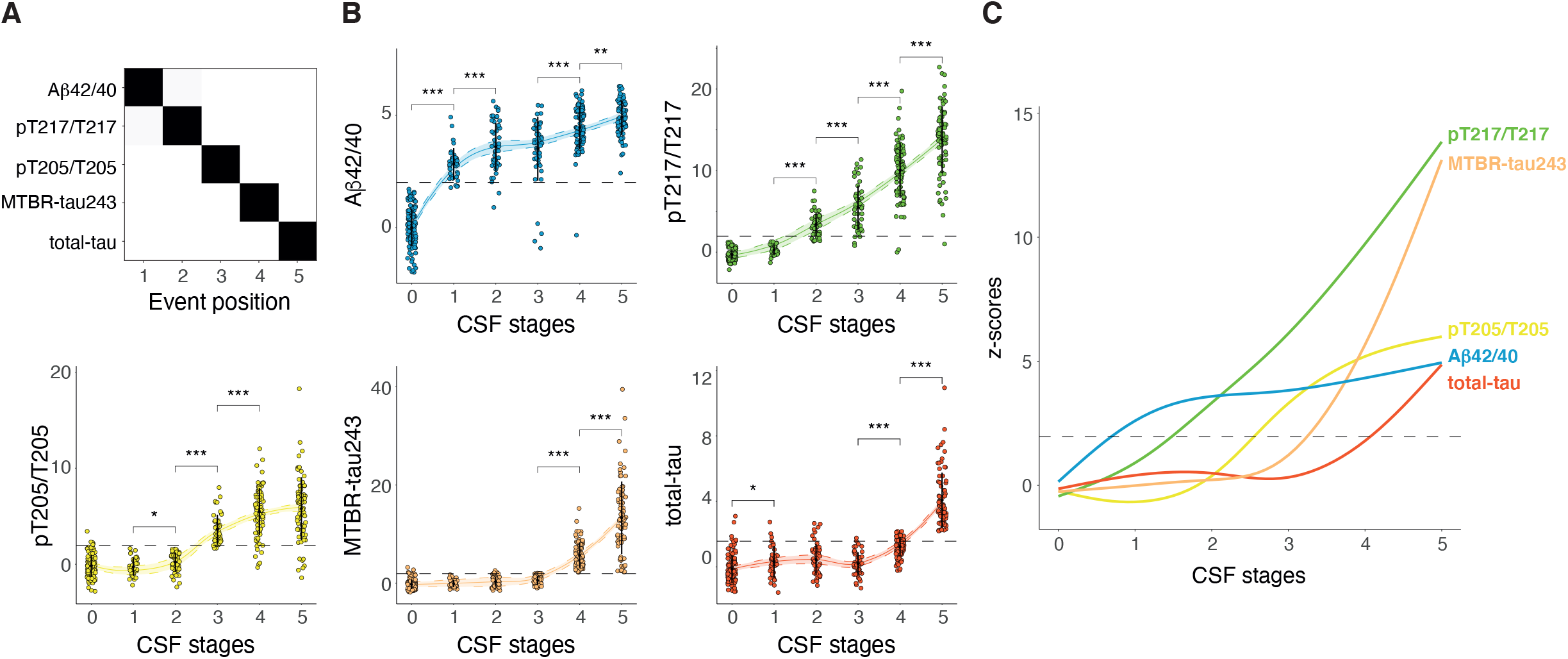
CSF staging model. Description of the CSF staging model and the levels of the biomarkers included in the model by CSF stage. Cross-validated confusion matrix of the CSF biomarkers of the model is shown in A. Biomarkers are sorted by the time they become abnormal based on the results of SuStaIn. Darkness represents the probability of that biomarker of becoming abnormal at that position, with black being 100%. Only amyloid-positive participants are included in this analysis. Individual biomarker levels by CSF stage in all BioFINDER-2 participants are shown in B. CSF levels are z-scored based on a group of CU− participants (n=63) and all increases represent increase in abnormality. Significant differences in contiguous CSF stages are shown with asterisks. Horizontal line is drawn at z-score=1.96 which represents 95%CI of the reference group (CU−). Smoothed LOESS lines of all CSF biomarkers are shown in C for comparison. CSF stage 0 represent being classified as normal by the model. Black dots and vertical lines represent mean and SD by CSF stage, respectively. *: p<0.05; **: p<0.01; ***: p<0.001. Abbreviations: Aβ, amyloid-β; CI, confidence interval; CU−, cognitively unimpaired amyloid negative; CSF, cerebrospinal fluid; LOESS, locally estimated scatterplot smoothing; MTBR, microtubule binding region; pT, phosphorylated tau; SuStaIn, subtype and stage inference.

We then examined the distribution of the CSF biomarkers included in the model by CSF biomarker stage. Individual plots by CSF biomarker can be found in Fig. 1B and statistics for each biomarker and their differences by CSF stage can be found in Supplementary Table 2. In summary, degree of abnormality of all biomarkers increased with higher CSF stages, although their trajectories were different. CSF Aβ42/40 and pT205/T205 had a steep increase at stage 1 and stage 3, respectively, and then continued increasing but in a lower degree. On the other hand, pT217/T217 (CSF stage 2) and MTBR-tau243 (CSF stage 4) levels continued to increase at all subsequent CSF stages in a similar degree after crossing the threshold for positivity. Also, both pT217/T217 and MTBR-tau243 reached very high levels compared to the reference control group (z-score>10). These different biomarker trajectories revealed that the included CSF biomarkers exhibit different behaviours across the disease *continuum*, aside from the biomarker disease stage at which they become abnormal. This is summarized in figure 1C, in which the smoothed locally estimated scatterplot smoothing (LOESS) regression of all CSF biomarkers are plotted.

Finally, we assessed the stability of our model using the longitudinal CSF data over a mean (SD) of 2.1 (0.2) years (n=220, Supplementary Table 1). We observed that most participants remained at the same stage (N=183, 83.2%) or progressed (n=29, 13.2%), while only few regressed (n=8, 2.9%, Ext Fig. 3A-B). Of those that progressed, most (n=25, 86.2%) progressed only one CSF stage during the two-year follow-up. This indicates a high stability of our model over time.

### CSF biomarker stages are associated with AD pathology, biomarkers and cognition

Next, we investigated the association between CSF stages and insoluble Aβ aggregates (Aβ-PET), insoluble tau aggregates (tau-PET), neurodegeneration (cortical thickness and CSF neurofilament light [NfL]) and cognition, using a global cognitive composite sensitive to early AD changes (modified version of preclinical Alzheimer’s Cognitive Composite [mPACC]I^35^, Fig. 2). The degree of biomarker abnormality increased with higher CSF stages, although the trajectories were different. For instance, Aβ-PET was the first to start increasing (already at CSF stage 1 [Aβ42/40 stage], Δz-score=1.12, p=0.032 compared to the previous stage) and continued to increase until the CSF stage 5 (total-tau stage), where it reached a plateau. However, it was at CSF stage 2 (pT217/T217) that the mean of Aβ-PET was above 1.96SD (95%CI) of CU− levels (mean[SD] z-score=4.01[2.71]). Tau-PET was the next biomarker to show a significant increase, emerging at CSF stage 3 (pT205/T205 stage, Δz-score=2.09, p<0.001), in which its mean was already above 1.96SD of CU− (mean[SD] z-score=2.02[3.16]). Notably, tau-PET levels continued to significantly increase in all subsequent CSF stages. The mPACC followed a similar trajectory, starting to increase also at CSF stage 3 (pT205/T205 stage, Δz-score=0.92, p=0.004), and crossing the 1.96SD threshold at the following stage (mean[SD] z-score=2.60[1.84]). Finally, both measures of neurodegeneration (cortical thickness and CSF NfL) showed a significantly lower degree of abnormality compared to the other biomarkers, even in the most advanced CSF stages. The abnormality of cortical thickness significantly increased at CSF stage 3 (pT205/T205 stage, Δz-score=0.80, p=0.006), but it was above the 1.96SD (95%CI) threshold only at the last CSF stage (total-tau stage, mean[SD] z-score=2.07[1.91]). CSF NfL, on the other hand, only showed significant differences between CSF stages 3 and 4 (Δz-score=0.36, p=0.016), but did not cross the 1.96SD threshold at any CSF stage. Statistics of each these AD biomarkers and their differences per CSF stage can be found in Supplementary Table 3.

**Fig 2:**
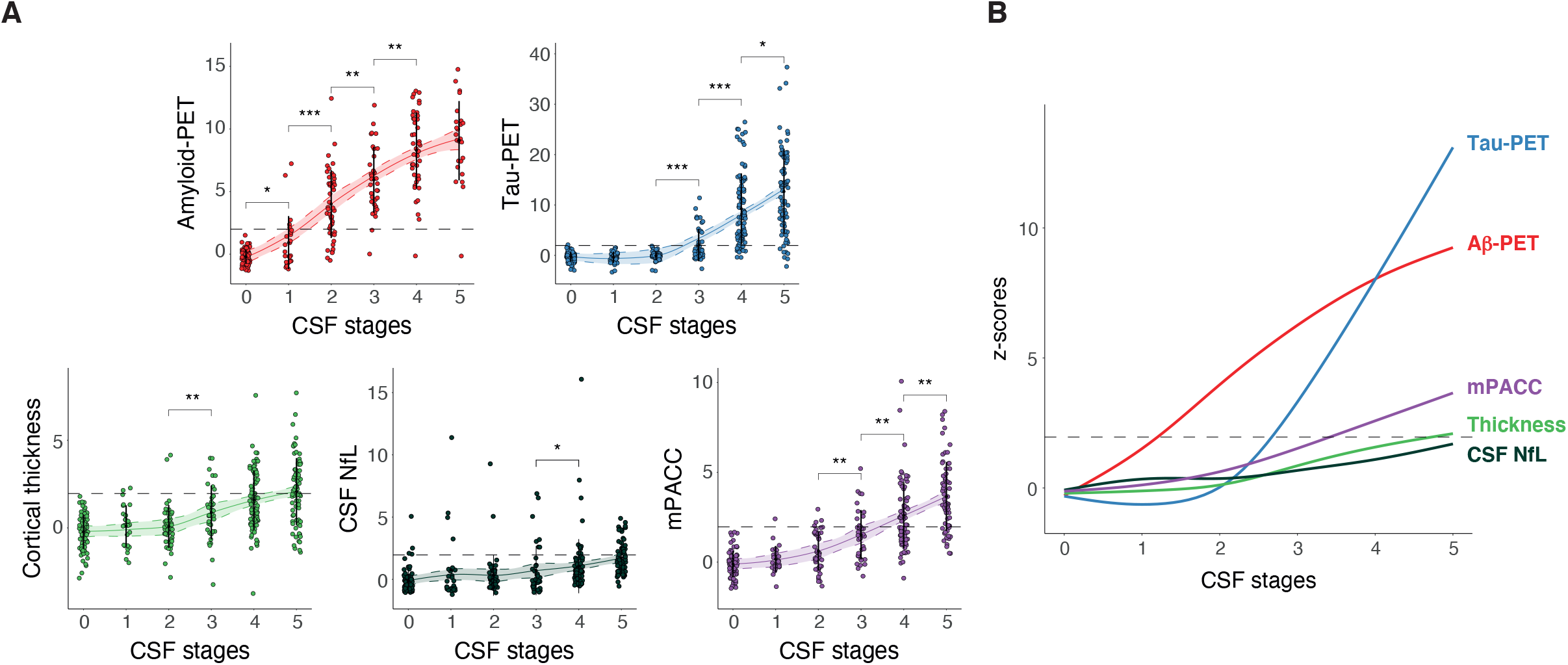
AD pathology, biomarkers and cognition by CSF stages. Depiction of individual biomarker levels, not used in the creation of the model, by CSF stage in BioFINDER-2 participants (A). These include biomarkers of amyloid (amyloid-PET) and tau (tau-PET in the meta-temporal ROI) pathologies, neurodegeneration (cortical thickness in the AD signature areas and CSF NfL) and cognition (mPACC). Biomarkers are z-scored based on a group of CU− participants (n=63) and all increases represent increase in abnormality. Significant differences in contiguous CSF stages are shown with asterisks. Horizontal line is drawn at z-score=1.96 which represents 95%CI of the reference group (CU−). Smoothed LOESS lines of all AD biomarkers are shown in B for comparison. All participants with available data were included in amyloid- and tau-PET analyses. For neurodegeneration (cortical thickness and NfL) and cognitive (mPACC) measures, we excluded non-AD dementia patients to avoid bias. Of note, only few AD dementia cases had amyloid-PET available due to study design. CSF stage 0 represent being classified as normal by the model. Black dots and vertical lines represent mean and SD per CSF stage, respectively. *: p<0.05; **: p<0.01; ***: p<0.001. Abbreviations: Aβ, amyloid-β; AD, Alzheimer’s disease; CI, confidence interval; CU−, cognitively unimpaired amyloid negative; CSF, cerebrospinal fluid; LOESS, locally estimated scatterplot smoothing; mPACC, modified preclinical Alzheimer’s cognitive composite; NfL, neurofilament light; PET, positron emission tomography; ROI, region of interest.

We further studied the associations between our CSF-based staging model and other biomarkers as additional analyses. For tau-PET, we quantified the signal in different brain regions, using the previously validated ROIs reflecting the different Braak stages^43^. Early (Braak I) and intermediate (Braak III-IV) regions of tau deposition become abnormal at CSF stage 3 (pT205/T205 stage, Braak I: mean z-score: 2.26; Braak III-IV: mean z-score: 1.97) and later regions (Braak V-VI) become abnormal at the CSF stage 4 (MTBR-tau243 stage, mean z-score: 4.58; Ext Data. Fig. 4 and Supplementary Table 4). Noteworthy, the number of participants with Braak I z-scores over 1.96SD at CSF stage 3 was higher than that with Braak III-IV (n=21, 42.9% vs. n=14, 28.6%).

We also examined different measures of cognitive function including composites for memory, executive, language and visuospatial functions, respectively. The first composite to cross the 1.96SD threshold was memory at CSF stage 4 (mean z-score: 2.18), and kept increasing in the next CSF stage (Ext. Data Fig. 5 and Supplementary Table 5). This was followed by the executive function composite, which was above the threshold at CSF stage 5 (t-tau stage, mean z-score: 2.15). Finally, the language and visuospatial cognitive composites showed an increasing degree of abnormality across CSF stages but did not cross the 1.96SD threshold at any CSF stage.

### CSF biomarker stages can be used for predicting A/T status and cognitive stages

Subsequently, we looked at the accuracy of our CSF staging model for predicting Aβ (A) and tau (T) status, as defined by PET^34^. We first looked at each independent pathology dichotomously (*i.e.*, positive or negative) and independently and, later, we looked at the ordinal categories merging both pathologies (*i.e.,* A-T-, A+T-, and A+T+). The number of positive participants by CSF stage and category are presented in Fig. 3A. Using receiver operating characteristic (ROC) curves analyses, we determined that CSF stage 2 (pT217/T217 stage) was the optimal threshold for predicting Aβ-PET positivity with high accuracy (area under the curve and 95% confidence interval [AUC[95%CI]]=0.96[0.93, 0.98], sensitivity=0.93 and specificity=0.89, first column Fig. 3B and Supplementary Table 6). Tau-PET positivity was also predicted with high accuracy when using CSF stage 4 (MTBR-tau243 stage) as a threshold (AUC[95%CI]=0.95[0.93, 0.97], sensitivity=0.91 and specificity=0.92, second column Fig. 3B and Supplementary Table 6).

**Fig 3:**
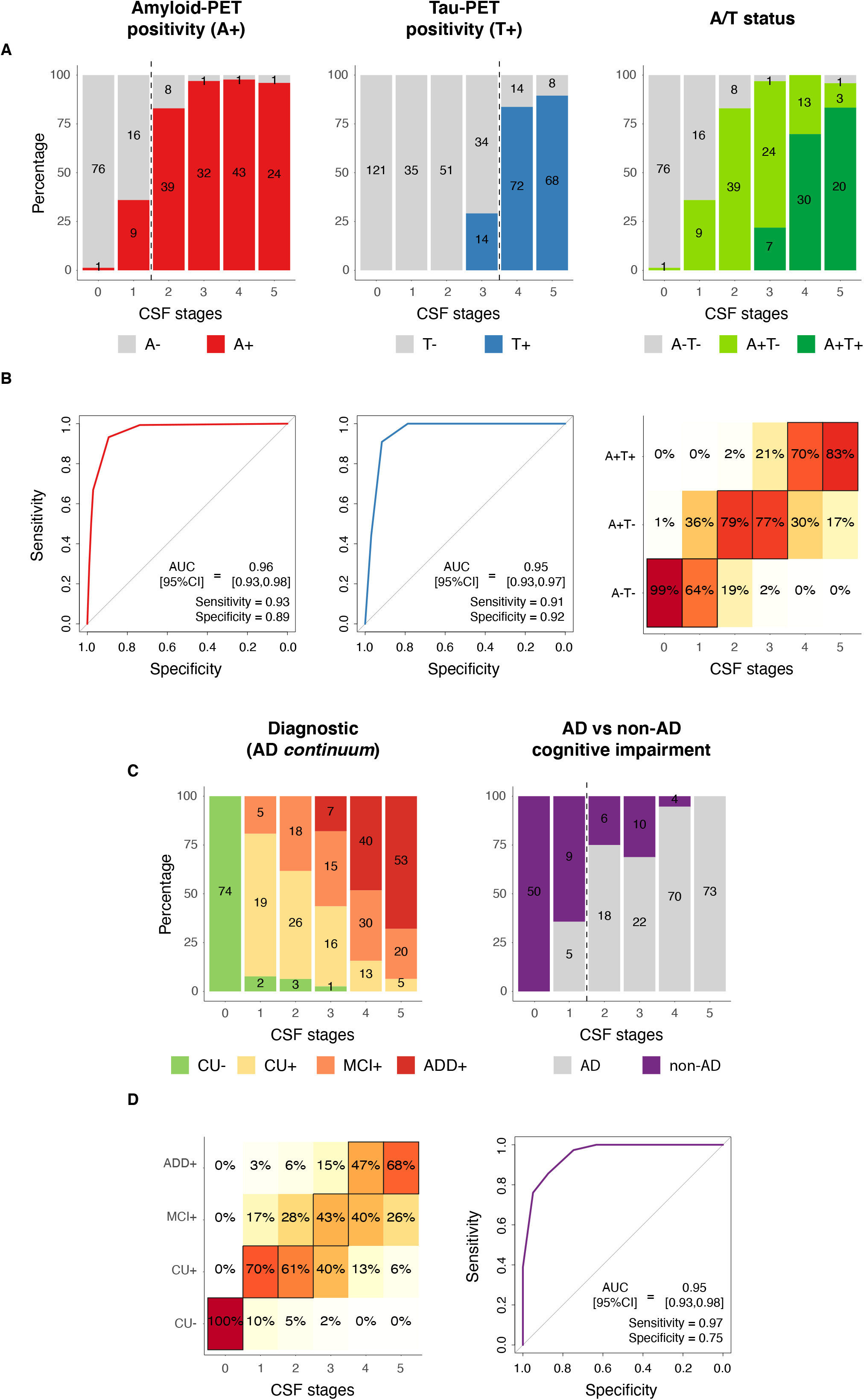
CSF stages for predicting A/T status and cognitive stages. CSF stages for predicting pathological status as measured with PET is shown in A-B, and for predicting cognitive stages and diagnostic groups in C-D. Barplots represent the number of participants in each category per CSF stage. Numbers of participants in each category per CSF stage are shown within the barplots (A and C). In B and D, ROC curves were used to assess the classification into dichotomic categories (Aβ-PET, tau-PET and AD vs non-AD cognitive impairment), whereas ordinal logistic regressions were used for ordinal categories (A/T status and diagnosis). Heatmaps represent the predicted percentage of participants in each outcome category (A/T or diagnosis) by CSF stage. The most probable (highest percentage) category by CSF stage is framed in black. For ROC analyses, AUCs, sensitivity and specificity measures from these analyses are shown in the plot. The optimal cut-off in each case is shown as a vertical dashed line in A or C. An A-T+ participant (n=1) was excluded from the A/T status analysis. Non-AD dementia cases were excluded from the cognitive stages analysis. And, only patients with objective impairment (MCI or dementia) were included in the analyses of AD vs. non-AD. Aβ- and tau-PET were assessed as positive based on previously validated cut-offs (Aβ: SUVR>1.03, tau: SUVR>1.36). Abbreviations: Aβ, amyloid-β; AD, Alzheimer’s disease; ADD+, Alzheimer’s disease dementia amyloid-positive; A-T-, amyloid-negative tau-negative; A-T+, amyloid-negative tau-positive; A+T-, amyloid-positive tau-negative; A+T+, amyloid-positive tau-positive; CSF, cerebrospinal fluid; CU−, cognitively unimpaired amyloid-negative; CU+, cognitively unimpaired amyloid-positive; MCI+, mild cognitive impairment amyloid-positive; PET, positron emission tomography; ROC, receiver operating characteristic; ROI, region of interest; SUVR, standardized uptake value ratio.

Ordinal logistic regression was used to assess the utility of CSF stages for predicting A/T status (*i.e.*, A-T-, A+T- or A+T+), and we calculated the c-index (an overall measure of discrimination equivalent to AUC for dichotomic outcomes) as a measure of accuracy. We observed that higher CSF stages were associated with higher predicted probabilities of being at more advanced A/T status (c-index[95%CI]=0.95[0.93, 0.97], last column Fig. 3B, and Supplementary Table 6). More specifically, participants at CSF stages 0 and 1 (negative biomarkers and Aβ42/40 stages) had the highest probability of being A-T-, at CSF stages 2 and 3 (pT217/T217 and pT205/T205 stages) being A+T- and at CSF stages 4 and 5 (MTBR-tau243 and total-tau stages) of being A+T+. Only one participant was classified as A-T+, which was excluded from this analysis.

Finally, we also aimed at investigating whether our staging model could be used as a diagnostic tool. In the first analysis, we used the CSF staging model for predicting cognitive stages within the AD continuum (*i.e.,* excluding non-AD). Higher CSF stages were associated with more advanced cognitive stages (c-index[95%CI]=0.88[0.86, 0.91], first column Fig. 3C and Supplementary Table 6). The model predicted that participants at CSF stage 0 (negative biomarkers stage) had the highest probability of being CU−; at CSF stages 1 and 2 (Aβ42/40 and pT217/T217 stages) were more probably CU+ (as assessed by CSF), at CSF stage 3 (pT205/T205) MCI+ and, finally, at CSF stages 4 and 5 (MTBR-tau243 and total-tau) ADD+. Lastly, we aimed at differentiating cognitive impairment due to AD or due to other neurodegenerative diseases. We therefore compared patients with AD to patients with non-AD dementia, only including those with objective cognitive impairment (*i.e.,* MCI and dementia patients). Participants at CSF stage 2 (pT217/T217 stage) or higher with objective cognitive impairment had a high probability of having AD as the cause of their cognitive impairment (AUC[95%CI]=0.95[0.93, 0.98], sensitivity=0.97 and specificity=0.75, last column Fig. 3C-D and Supplementary Table 6).

### Longitudinal rates of change of AD biomarkers differ by CSF stages

Next, we used longitudinal imaging and cognitive data to assess how AD biomarkers change over time based on the baseline CSF stage classification (Supplementary Table 7). The rate of accumulation of Aβ aggregates as measured with PET (n=218) increased at early CSF stages reaching the highest values at CSF stage 2 (pT217/T217 stage) and thereafter the rate decreased but still remained positive (Fig. 4 and Supplementary Table 8). On the other hand, the tau-PET (n=312), cortical thickness (n=300) and mPACC (n=342) exhibited monotonic increases in rates of change over time, with the rates starting to be significantly different from contiguous CSF stages at CSF stage 3 (pT205/T205 stage; Fig. 4). Figure 4B depicts that tau-PET, followed by mPACC had the highest rate of change (z-scored), while Aβ-PET and cortical thickness had lower rates of change that were in a similar range.

**Fig 4:**
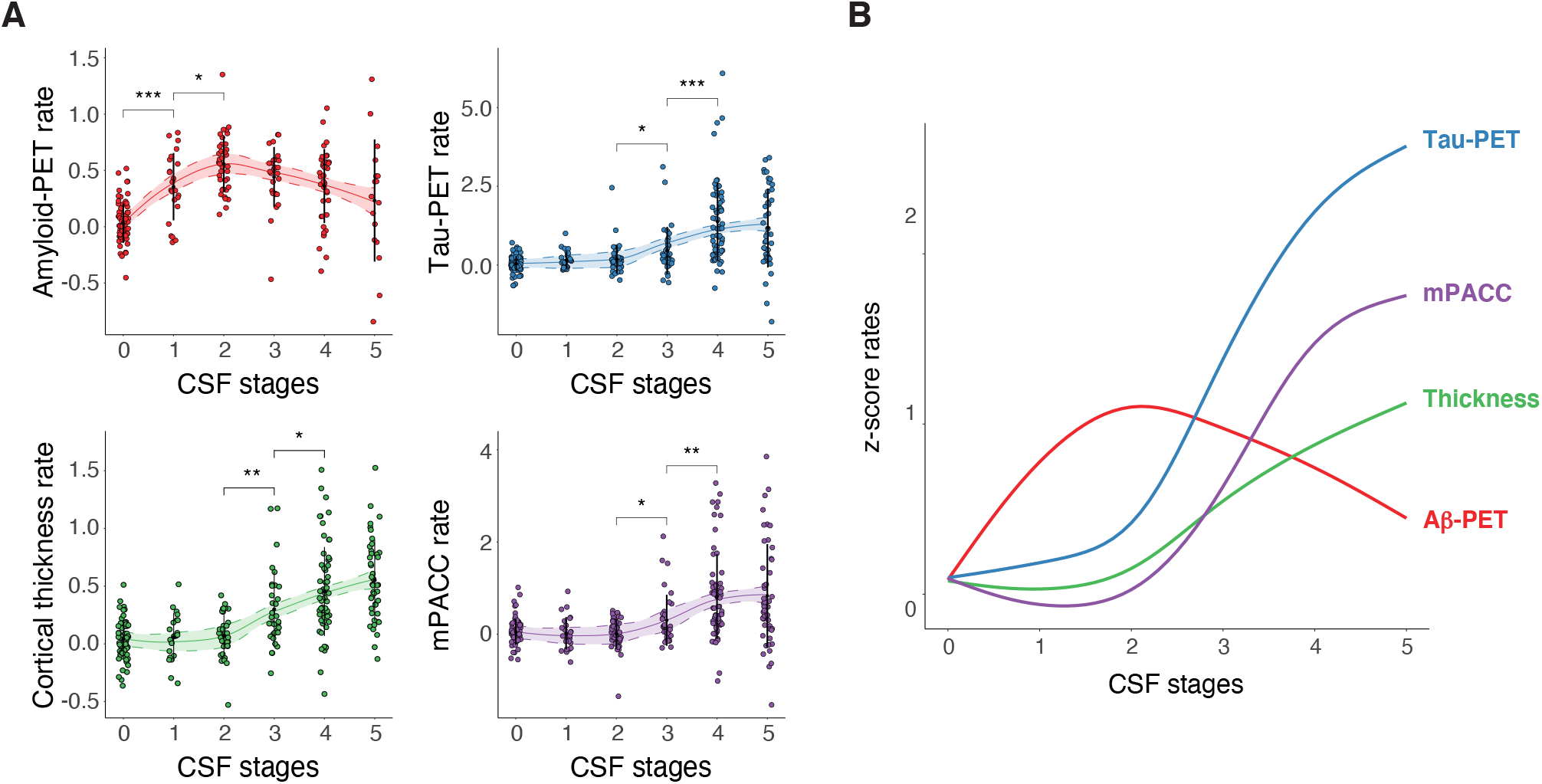
Longitudinal rate of change of AD biomarkers by CSF stages. Depiction of individual biomarker longitudinal rates of change by CSF stage in BioFINDER-2 participants (A). These include biomarkers of amyloid (amyloid-PET) and tau (tau-PET in the meta-temporal ROI) pathologies, neurodegeneration (cortical thickness in the AD signature) and cognition (mPACC). Biomarkers are z-scored based on a group of CU− participants (n=63) and all increases represent increase in abnormality. Rates of change were calculated with individual linear regression models. Significant differences in contiguous CSF stages are shown with asterisks. Smoothed LOESS lines of all AD biomarkers are shown in B for comparison. All participants were included in amyloid- and tau-PET analyses. For neurodegeneration (cortical thickness) and cognitive (MMSE) measures, we excluded non-AD dementia patients to avoid bias. CSF stage 0 represent being classified as normal by the model. Black dots and vertical lines represent mean and SD per CSF stage, respectively. *: p<0.05; **: p<0.01; ***: p<0.001. Abbreviations: Aβ, amyloid-β; AD, Alzheimer’s disease; CI, confidence interval; CU−, cognitively unimpaired amyloid negative; CSF, cerebrospinal fluid; LOESS, locally estimated scatterplot smoothing; mPAC, modified preclinical Alzheimer’s cognitive composite; PET, positron emission tomography; ROI, region of interest.

### CSF biomarker stages predict clinical progression

In the next set of analyses, we tested whether our CSF staging model was useful for predicting subsequent clinical progression (up to 5-years of follow-up after the baseline visit). First, we tested the ability of our model to predict progression to AD dementia from CU or MCI status at baseline (progressors: n=41). Based on Kaplan-Meier analyses (Fig. 5A), participants at higher CSF stages (4-5; MTBR-tau243 and total-tau stages) at baseline had higher probability to progress to AD dementia, than those at positive lower CSF stages (*i.e.*, 1-3). When adjusting for age, sex, and clinical status at baseline (*i.e.*, CU or MCI), the hazard ratio (HR) was 5.2 (95%CI: [2.2, 12.6], p<0.001) when comparing participants at CSF stages 4 or 5 to participants at lower, but positive, CSF stages (1-3, Aβ42/40 to pT205/T205, reference; Fig. 5B and Supplementary Table 9). When including only those with MCI at baseline (progressors: 38/88), we still found that those at CSF stages 4 or 5 at baseline had a significantly higher probability to progress to AD dementia (HR[95%CI]=4.5[1.8, 10.8], p<0.001, Fig. 5C-D and Supplementary Table 9). Finally, we investigated the utility of the CSF staging model when predicting progression from CU to MCI status (progressors: 11/159). Again, those CU participants at higher CSF stages (4-5) at baseline, were much more prone to progress to MCI with a HR of 16.0 (95%CI: 3.2, 81.1, p<0.001, Fig. 5E-F and Supplementary Table 9) compared to those in stage 1-3, supporting the clinical utility of the proposed staging model. There were no progressors from CSF stage 0 in any case, which prevented us from comparing these participants with the other CSF stages groups. Kaplan-Meier curves for each individual CSF stage are depicted in Ext Data Fig. 6.

**Fig 5:**
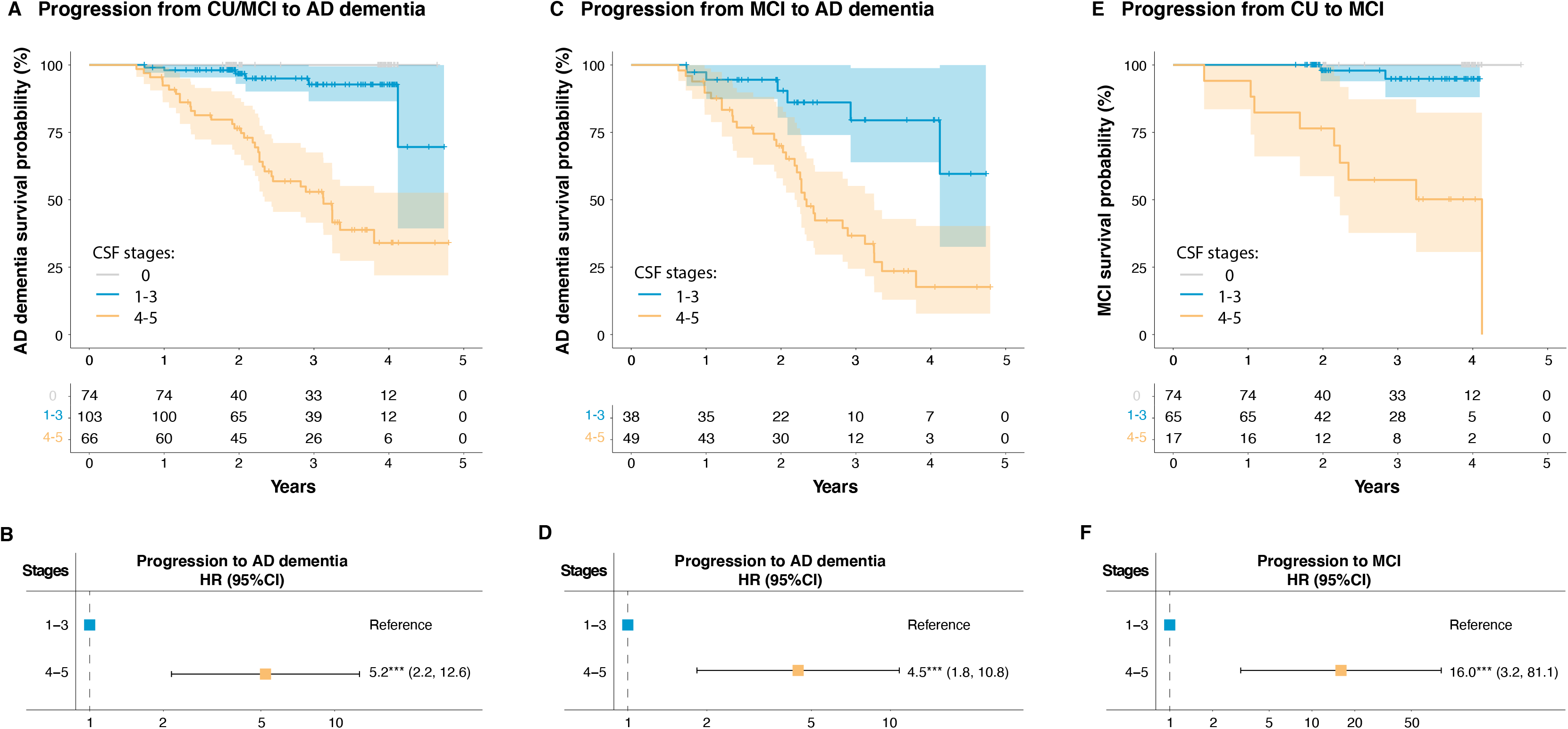
CSF stages for predicting clinical progression. Higher CSF stages groups (4-5) show higher HR of clinical progression compared to lower positive stages (reference: 1-3). Progression from CU or MCI at baseline to AD dementia is shown in A-B. Progression from MCI at baseline to AD dementia is shown in C-D. Progression from CU at baseline to MCI is shown in E-F. Kaplan-Meier curves, as well as the number of participants per group and timepoint are shown in A, C and E, respectively. Cox-proportional hazards models were used to calculate HR[95%CI] of higher CSF stages (4-5) compared to the reference (1-3, B, D and F). These analyses were adjusted for age, sex in all cases, and additionally for clinical status at baseline (CU or MCI) if appropriate. Abbreviations: AD, Alzheimer’s disease; CI, confidence interval; CSF, cerebrospinal fluid; HR, hazard ratios; MCI, mild cognitive impairment.

### Replication in an independent cohort

Finally, we replicated the staging model and the main analyses in the Charles F. and Joanne Knight Alzheimer’s Disease Research Center (Knight-ADRC) cohort (n=222, Table 2). SuStaIn selected one unique subtype as the optimal model with the same CSF abnormality ordering as the one previously obtained in BioFINDER-2 (Fig. 6A). In this cohort, however, there was slightly higher uncertainty between the ordering of first two (Aβ42/40 and pT217/T217) and the last two (MTBR-tau243 and total-tau) stages. These differences may be mostly due to the difference in sample size, especially in more advanced AD cases (only 9 mild AD dementia cases). Nonetheless, the overall behaviour of these CSF biomarkers by the biomarker stages was similar to that in the main cohort (Fig. 6B and Supplementary Table 2). Further, the other AD biomarkers available (not included in the CSF staging model), showed similar trajectories to those in the discovery (BioFINDER-2) sample. Aβ-PET was once again the first modality to cross the 1.96SD threshold at CSF stage 1 (Aβ42/Ab40 stage), followed by tau-PET in CSF stage 3 (pT205/T205 stage) and, finally, CSF NfL crossing this threshold at the final stage (total-tau stage; Fig. 6C and Supplementary Table 3). Neither the global cognitive composite nor AD cortical thickness signature crossed this threshold. The main difference compared with BioFINDER-2, was the lower degree of abnormality for all markers in the last CSF stages. This might be explained by the lower number of advanced patient cases in this cohort. The individual plots for each CSF and imaging biomarker by CSF stages are shown in Ext Data Fig. 7. Details of participants characteristics (Ext Data Fig. 2), tau-PET binding in different regions (Ext Data Figure 4 and Supplementary Table 4) and other cognitive measures (Ext Data Figure 5 and Supplementary Table 5) per CSF stage can be found in the Ext Data. Stability analyses, within participants with available longitudinal CSF measures (n=51, Supplementary Table 10), also showed that most participants remained at the same stage (n=46, 90.2%) or progressed (n=4, 7.8%) at follow-up (Ext Data Fig. 3C-D).

**Table 2:** Knight-ADRC participants’ characteristics. Data is shown as mean(SD) unless otherwise stated. Participants are divided by clinical diagnosis and amyloid status based on their CSF Aβ42/40 levels (Aβ+: <0.0673). Very mild AD dementia patients had a CDR=0.5 and mild AD dementia patients had a CDR≥1, both with AD as etiology. Other dementias group includes participants with CDR>0 with non-AD etiology. Only participants who progressed to CDR≥0.5 or CDR≥1 due to AD etiology were considered to progress. ^a^, 1 participant missing; ^b^, 3 participants missing; ^c^, 5 participants missing; ^d^, 2 participants missing; ^e^, 4 participants missing; ^f^, 8 participants missing. Abbreviations: Aβ, amyloid-β; AD, Alzheimer’s disease; ADD+, Alzheimer’s disease dementia amyloid positive; CU−, cognitively unimpaired amyloid negative; CU+, cognitively unimpaired amyloid positive; CSF, cerebrospinal fluid; MCI, mild cognitive impairment amyloid positive; mPACC, modified preclinical Alzheimer’s cognitive composite; nonAD, non-Alzheimer’s related disease; PET, positron emission tomography; ROI, region of interest; SD, standard deviation.

|  | All<br>(n=222) | CU-<br>(n=84) | CU+<br>(n=98) | Very mild<br>AD (n=24) | AD<br>dementia<br>(n=9) | Other<br>dementias<br>(n=7) |
| --- | --- | --- | --- | --- | --- | --- |
| Age, years | 71.2 (7.7) | 67.6 (7.2) | 73.2 (7.4) | 73.4 (6.4) | 74.1 (7.6) | 75.0 (7.2) |
| Women, n(%) | 112<br>(50.5%) | 39<br>(46.4%) | 56<br>(57.1%) | 9<br>(37.5%) | 4<br>(44.4%) | 4<br>(57.1%) |
| APOE-ε4<br>carriership,<br>n(%)<sup>a</sup> | 99 (44.6%) | 18 (21.4%) | 56 (57.1%) | 15 (62.5%) | 6 (66.7%) | 4 (57.1%) |
| Years of<br>education | 16.3 (2.5) | 16.5 (2.3) | 16.5 (2.4) | 14.9 (2.8) | 14.7 (2.6) | 17.7 (2.2) |
| Amyloid-PET,<br>Centiloids | 44.0 (41.2) | 7.6 (11.2) | 57.7 (33.3) | 83.6 (23.7) | 119 (38.3) | 57.9 (46.8) |
| Tau-PET,<br>SUVR<sup>b</sup> | 1.24 (0.22) | 1.13 (0.08) | 1.22 (0.13) | 1.51 (0.38) | 1.72 (0.25) | 1.23 (0.12) |
| Cortical<br>thickness,<br>mm | 2.52 (0.16) | 2.59 (0.12) | 2.53 (0.14) | 2.39 (0.17) | 2.29 (0.15) | 2.35 (0.19) |
| CSF NfL<sup>c</sup> | 1000 (578) | 740 (313) | 1010 (489) | 1660 (974) | 1360 (404) | 1230 (569) |
| Global<br>cognitive<br>composite, z-score<sup>d</sup> | 0.44<br>(1.11) | -0.01<br>(0.72) | 0.36<br>(0.75) | 2.01<br>(0.96) | 3.86<br>(2.36) | 1.12<br>(1.40) |
| Progressed<br>to CDR≥0.5<sup>e</sup> | 41 (18.5%) | 8 (9.5%) | 33 (33.7%) | - | - | - |
| Progressed<br>to CDR≥1<sup>f</sup> | 30 (14.5%) | 0 (0%) | 14 (14.3%) | 16 (66.7%) | - | - |

We also calculated the optimal CSF stages for predicting Aβ-PET and tau-PET positivity using ROC curves. As in the case of BioFINDER-2, CSF stage 2 (pT217/T217 stage) was optimal for predicting Aβ-PET positivity (AUC[95%CI]=0.89[0.85, 0.94], Fig. 6D&G and Supplementary Table 6), whereas CSF stage 4 (MTBR-tau243 stage) was optimal for predicting tau-PET positivity (AUC[95%CI]=0.94[0.91, 0.96], Fig. 6E&H). Consistent with findings in BioFINDER-2, higher CSF stages were predictive of more advanced A/T stages, as assessed by PET (c-index[95%CI]=0.89[0.86, 0.92], Fig. 6 F&I, Supplementary Table 6). Being at CSF stages 0 and 1 (biomarker negative and Aβ42/40 stage) was highly predictive of being A-T-, being at CSF stages 2 and 3 (pT217/T217 and pT205/T205 stages) was predictive of being A+T- and, at CSF stages 4 and 5 (MTBR-tau243 and total-tau stages) of being A+T+.

**Fig 6:**
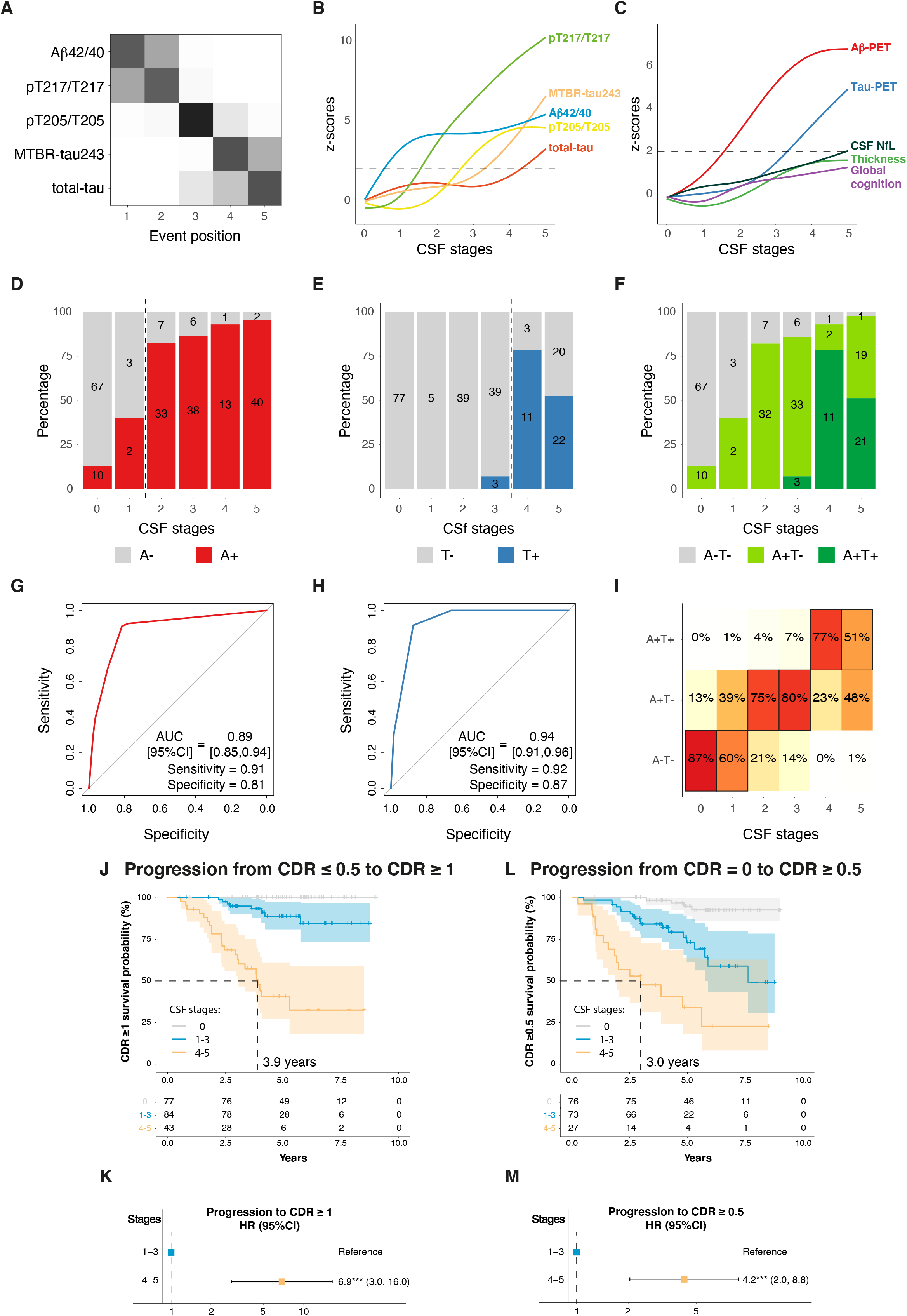
Replication of main analyses in Knight-ADRC participants. Description of the model is shown in A-B. Cross-validated confusion matrix of the CSF biomarkers of the model is shown in A. Biomarkers are sorted by the time they become abnormal based on the results of SuStaIn. Darkness represents the probability of that biomarker of becoming abnormal at that position, with black being 100%. Only amyloid-positive participants are included in this analysis. Description of the CSF levels of the biomarkers included in the model by CSF stage are shown in B for all Knight-ADRC participants. Depiction of individual biomarker levels, not used in the creation of the model, by CSF stage are shown in C. These include biomarkers of amyloid (amyloid-PET) and tau (tau-PET in the meta-temporal ROI) pathologies, neurodegeneration (cortical thickness in the AD signature areas and CSF NfL) and cognition (global cognitive composite). CSF and AD biomarker levels are z-scored based on a group of CU− participants (n=71) and all increases represent increase in abnormality. Horizontal line is drawn at z-score=1.96 which represents 95%CI of the reference group (CU−). CSF stage 0 represent being classified as normal by the model. Prediction of amyloid-PET (D-G), tau-PET (E-H) and A/T status (by PET, F-I) are shown next. Number of participants in each category are colored in D, E and F. Numbers of participants in each category per CSF stage are shown within the barplots. In G and H, ROC curves were used to determine the CSF stage to optimally classify participants into positive/negative in each case. AUCs, sensitivity and specificity measures from these analyses are shown in the plot. The optimal cut-off in each case is shown as a vertical dashed line in D and E, respectively. Ordinal logistic regression was used for assessing A/T status (I). The heatmap represent the predicted percentage of participants in each A/T group per CSF stage. The most probable (highest percentage) group per CSF stage is framed in black. Amyloid-PET was considered positive if Centiloids>20, tau-PET was considered positive if SUVR at meta-temporal ROI was higher than 1.32. An A-T+ participant (n=1) was excluded from the A/T status analysis. Higher CSF stages groups (4-5) show higher HR of clinical progression compared to lower stages (reference: 1-3, J-M). Progression from CDR=0 or CDR=0.5 at baseline to CDR≥1 is shown in J-K. Progression from CDR=0 at baseline to CDR≥0.5 is shown in L-M. Kaplan-Meier curves, as well as the number of participants per group and timepoint are shown in J and L. Cox-proportional hazards models were used to calculate HR[95%CI] of higher CSF stages (4-5) compared to the reference (1-3, K and M). These analyses were adjusted for age, sex in all cases, and additionally for clinical status at baseline (CDR=0 or CDR=1) if appropriate. Abbreviations: Aβ, amyloid-β; AD, Alzheimer’s disease; AUC, area under the curve; CDR, clinical dementia rating; CI, confidence interval; CU−, cognitively unimpaired amyloid negative; CSF, cerebrospinal fluid; HR, hazard ratio; LOESS, locally estimated scatterplot smoothing; MMSE, Mini-mental state examination; MTBR, microtubule binding region; NfL, neurofilament light; PET, positron emission tomography; ROC, receiver operating characteristic; SuStaIn, subtype and stage inference; SUVR, standardized uptake value ratio.

Finally, we investigated the prognostic capacity of our model for predicting progression to CDR≥1 (AD dementia, progressors: 41/218) and CDR≥0.5 (MCI or very mild AD dementia, progressors: 30/214). We found that CU (CDR=0) and very mild AD (CDR=0.5) participants at the highest CSF stages (4-5; MTBR-tau243 and total-tau stages) exhibited an increased risk (HR[95%CI]=6.9[3.0, 16.0], p<0.001) of progressing to AD dementia (CDR≥1) at follow-up, even when adjusting for age, sex and clinical status (*i.e.*, CDR=0 or CDR=0.5) at baseline, compared to participants at CSF stages 1-3 (Fig. 6J-K and Supplementary Table 9). Similarly, CU participants at higher CSF stages (*i.e.*, 4-5) had higher risk (HR[95%CI]=4.2[2.0, 8.8], p<0.001) of progressing to very mild AD or more advanced disease stages when compared to participants at lower, but positive, CSF stages (1-3, Fig. 6L-M and Supplementary Table 9). In this case, participants at CSF stages 1-3 also showed significant higher risk to progress to CDR≥0.5 than those at CSF stage 0 (HR[95%CI]=5.0[1.6, 15.0], p=0.005). There were no progressors to CDR≥1 at CSF stage 0, which prevented us to compare this group to the others. Kaplan-Meier curves for each individual CSF stage are depicted in Ext Data Fig. 6.

## Discussion

In this study, we created and evaluated a staging model for AD using five CSF biomarkers reflecting abnormalities of soluble Aβ and different soluble tau species. We have demonstrated that a single CSF collection is sufficient to accurately stage participants representing the entire AD *continuum*. This is possible because CSF biomarker abnormalities followed a stereotypical trajectory in all participants, which enabled a single staging model usable for everyone. Notably, we have been able to relate the CSF stages of our model to abnormality in other well-described AD biomarkers, such as Aβ-PET, tau-PET, MRI and in cognitive measures. Further, our CSF staging model was able to accurately predict positivity of the imaging biomarkers of Aβ and tau, and to predict A/T status, as assessed by PET. The CSF staging model was also related to cognitive stages and was able to differentiate cognitive impairment due to AD from other dementias. Importantly, we also observed different longitudinal rates of change of AD biomarkers at different CSF stages, which may allow us to determine which participants will progress more in key aspects of the disease. We also showed that participants in the more advanced stages of our CSF-based model were at higher risk for clinical decline. And, finally, we were able to replicate the model and main results in an independent cohort. Altogether, these results prove the validity and clinical utility of our CSF staging model, suggesting that it may hold promise for both clinical practice and in clinical trials^44^.

The first aim of this analysis sought to establish whether there was a stereotypic ordering in when key CSF biomarkers become abnormal. SuStaIn is an optimal approach to solve this question, as it allows the modelling of different trajectories, if existent, using cross-sectional data^41^, as has been successfully applied to imaging biomarkers^35,36,45^. We observed that the CSF biomarkers investigated in this study became abnormal in a particular sequence, and more importantly, that this sequence did not vary systematically across participants. This result is important by itself as it tells us that there may be a single cascade of events that leads to sequential abnormality of these soluble proteins in the brain, common to all AD patients. Previous studies already suggested that changes in the levels of tau fragments phosphorylated at different sites may be linked mechanistically and could be associated with disease stages^46–50^. Based on our results, Aβ plaques reflected by an imbalance of soluble amyloid species (*i.e.*, low Aβ42/40) may drive hyper-phosphorylation of tau in early phosphorylation site (pT217/T217), as previously suggested by human and animal data^51,52^, which would subsequently be followed by hyper-phosphorylation in later site (pT205/T205) and eventually increase other tau fragments (MTBR-tau243 and total tau) due to tangles formation and neurodegeneration, respectively. Notably, this sequence of events is in line with previous literature^53,54^, and demonstrates that late onset sporadic AD molecular pathway matches the same sequence of events as autosomal dominant AD^29^. Exploring in detail this cascade of events may provide mechanistic insights into disease pathology and progression. And, in turn, could have important consequences in drug development, as targeting some of the earliest events of this sequence may stop or reduce subsequent events in the cascade and thereby having a significant effect on tau aggregation^46,50,55^.

Nonetheless, perhaps the most important result of our study was proving the utility of a CSF model as a method to stage AD *in vivo*^44^. In our model, CSF stages could be related to main molecular changes and clinical tipping points in the course of the disease, including abnormal levels of deposited Aβ (CSF stage 2: pT217/T217)^19,20,23,25,26,28,29,32^ and tau (CSF stages 3: pT205/T205^28,29,32^), early cognitive impairment (CSF stage 4: MTBR-tau243^32^) and neurodegeneration (CSF stage 5: total tau^42^), following the expected pattern. With the objective of characterizing the molecular status of the participants using our model, we observed that participants at CSF stages 2 and 3 (pT217/T217 and pT205/T205 stages) could be categorized with high accuracy as being Aβ-positive and tau-negative by PET (A+T-), while participants at CSF stage 4 (MTBR-tau243) or higher were Aβ- and tau-PET positive (A+T+)^42^. Importantly, these cut-points were reproduced in the Knight-ADRC cohort, even using different PET tracers and quantification methods supporting the consistency of the model. Being able to accurately assess Aβ and tau status with a single CSF collection may be very useful to select the optimal participants for a clinical trial, such as has been done in the donanemab trial (NCT03367403)^11^, without the need of acquiring both an Aβ-PET and tau-PET scan to determine if a patient is eligible for treatment. In BioFINDER-2, we also observed the diagnostic utility of this CSF staging model, as it was able to accurately discern AD from non-AD related cognitive impairment, and could differentiate cognitive stages. Thus, the use of our model as a diagnostic tool may have important consequences also at the clinical level.

Notably, our CSF staging model also showed prognostic utility. First, we observed that participants at different CSF stages showed different rates of change in multiple biomarkers. For instance, rates of Aβ accumulation across CSF stages showed the previously reported inverted U shape^3,56^, with participants at CSF stage 2 (pT217/T217) exhibiting the highest rates of change. On the other hand, the other imaging biomarkers and cognitive scores showed increased rate of change with increasing CSF stages, only plateauing at the last stage, as expected^57^. These results support the use of our staging model as an enrichment technique for clinical trials^58^. But, more importantly, we also observed that the CSF staging model was able to predict clinical progression. Being at the later stages of our model, increased the risk of progressing to AD dementia, even when accounting for cognitive status at baseline (Fig. 5). Further, we also observed a higher risk of progressing to MCI or very mild AD, although this analysis should be replicated in larger cohorts with longer follow-up. Notably, the prognostic ability of our CSF staging model was replicated in the Knight-ADRC cohort. These results suggest a clear prognostic utility on staging participants based on their CSF profile, which may imply significant reductions in costs and complexity compared to previous staging methods based on PET^34,36,37,59^.

Importantly, we view the present model as a first step toward providing meaningful disease progression staging using a single CSF measurement^44^. We expect that additional biomarkers will be included to the model to either gain further granularity in specific disease stages, or to signify to other pathophysiological events (*e.g.*, microglial reactivity)^60^. Being able to measure several pathophysiological abnormalities using one sample is one of the main advantages of using fluid samples instead of PET for staging. Another advantage of this model is that financial and infrastructure cost of CSF is low compared to other measures, such as PET. Looking toward the future, we hope to be able translate these results into plasma biomarkers, which would facilitate even greater availability and cost-effectiveness. Widespread use of our fluid biomarker staging model in primary care would likely require replacing CSF measures with plasma measures without greatly sacrificing model performance. Efforts in this direction are currently underway, but development of reliable plasma assays for pT205/T205 and MTBR-tau243 are still ongoing.

This study has several strengths but also some limitations. The main strength of this study is the proven utility of the model, which was replicated in an independent cohort, and thereby supports the generalizability of our staging model. Another important strength is the use of several biomarkers measured with very high-performing assays^28,61^, which is crucial for the accurate assessment of pathology^4^. However, some limitations must be recognized. Although we included CSF biomarkers with proven utility, we acknowledge that there are some other interesting markers, such as p-tau235^62^, that have not been analyzed in this study. However, we think that our CSF staging model in its current form was still successful at signaling the main inflection points of the disease. Further, p-tau231, which is thought to become abnormal early in the disease^20–22^, although not always^25,61^, was excluded from our model as it followed a similar abnormality tendency as pT217/T217, without providing better performance for staging than the later. We hypothesize that this may be in part related to difference in analytical performances, as the mass spectrometry platform used in our study provided rather higher coefficient of variation for pT231/T231 measurements (12-18% compared to 5-7% for pT217/T217). Future studies in earlier cohorts or with optimized assays for measuring p-tau231 should test whether the present model could be improved. Another important issue is that we acknowledge that CSF collection require trained clinicians, and we plan to move towards a plasma-based staging models when these biomarkers become available. A replication of these results in a more diverse population is also needed to confirm the utility of our model in a less selected population. We would also like to point out that the CSF stages proposed here are related to events of the disease and not to time. Thus, it may be possible that the time for progressing from one CSF stage to the next vary significantly depending on the CSF stage at baseline.

In conclusion, in this study we have developed an accurate staging model for AD based on only five CSF biomarkers, and we have evaluated it in two large independent cohorts. We have shown that the model is stable, and accurately reflects biomarker changes in AD, providing an easier and cheaper method for characterization of participants for both clinical setting and trials. Further, our model has demonstrated its utility for prognosis, being able to identify participants with more pronounced longitudinal changes in AD biomarkers as well as those individuals with higher risk of deteriorating in cognitive status. This CSF staging model may be a useful, cheap, and accessible method in clinical trials for optimal selection of study participants and as a surrogate outcome measure. Further, the staging model has great potential for use in clinical practice in the diagnostic and prognostic work-up of patients with cognitive symptoms and potentially also for selecting optimal candidates for disease-modifying treatments. And we expect it may have an influence on the update of the A/T/(N) criteria.^42^ Altogether, our staging model may be an important step towards a more sophisticated personalized medicine approach of AD, which will be key with the advancement of novel disease-modifying treatments.

## DATA AVAILABILITY

The datasets generated and/or analyzed during the current study are available from the authors (O.H and R.J.B). We will share datasets within the restrictions of IRB ethics approvals, upon reasonable request.

For BioFINDER-2 data, anonymized data will be shared by request from a qualified academic investigator for the sole purpose of replicating procedures and results presented in the article and as long as data transfer is in agreement with EU legislation on the general data protection regulation and decisions by the Ethical Review Board of Sweden and Region Skåne, which should be regulated in a material transfer agreement.

## DISCLOSURES

OH has acquired research support (for the institution) from ADx, AVID Radiopharmaceuticals, Biogen, Eli Lilly, Eisai, Fujirebio, GE Healthcare, Pfizer, and Roche. In the past 2 years, he has received consultancy/speaker fees from AC Immune, Amylyx, Alzpath, BioArctic, Biogen, Cerveau, Eisai, Eli Lilly, Fujirebio, Genentech, Merck, Novartis, Novo Nordisk, Roche, Sanofi and Siemens. JWV is supported by the SciLifeLab & Wallenberg Data Driven Life Science Program (grant: KAW 2020.0239). KH is an Eisai-sponsored voluntary research associate professor at Washington University and has received salary from Eisai. Washington University, RJB, and DMH have equity ownership interest in C2N Diagnostics. RJB. and DMH receive income from C2N Diagnostics for serving on the scientific advisory board. KH, NRB, and RJB. may receive income based on technology (METHODS TO DETECT MTBR TAU ISOFORMS AND USE THEREOF) licensed by Washington University to C2N Diagnostics. DMH may receive income based on technology (ANTIBODIES TO MID-DOMAIN OF TAU) licensed by Washington University to C2N Diagnostics. RJB has received honoraria as a speaker, consultant, or advisory board member from Amgen and Roche. DMH is on the scientific advisory board of Genentech, Denali, and Cajal Neurosciences, and consults for Alector and Asteroid. NRB is co-inventor on the following US patent applications: ‘Methods to detect novel tau species in CSF and use thereof to track tau neuropathology in Alzheimer’s disease and other tauopathies’ (PCT/US2020/046224); ‘CSF phosphorylated tau and amyloid beta profiles as biomarkers of tauopathies’ (PCT/US2022/022906); and ‘Methods of diagnosing and treating based on site-specific tau phosphorylation’ (PCT/US2019/030725). NRB may receive a royalty income based on technology licensed by Washington University to C2N Diagnostics.

## FUNDING

The BioFINDER study was supported by the Swedish Research Council (2022-00775), ERA PerMed (ERAPERMED2021-184), the Knut and Alice Wallenberg foundation (2017-0383), the Strategic Research Area MultiPark (Multidisciplinary Research in Parkinson’s disease) at Lund University, the Swedish Alzheimer Foundation (AF-980907), the Swedish Brain Foundation (FO2021-0293), The Parkinson foundation of Sweden (1412/22), the Cure Alzheimer’s fund, the Konung Gustaf V:s och Drottning Victorias Frimurarestiftelse, the Skåne University Hospital Foundation (2020-O000028), Regionalt Forskningsstöd (2022-1259) and the Swedish federal government under the ALF agreement (2022-Projekt0080). The precursor of ^18^F-flutemetamol was sponsored by GE Healthcare. The precursor of ^18^F-RO948 was provided by Roche. GS received funding from the European Union’s Horizon 2020 research and innovation program under the Marie Sklodowska-Curie action grant agreement No 101061836, from an Alzheimer’s Association Research Fellowship (#AARF-22-972612), Greta och Johan Kocks research grants and, travel grants from the Strategic Research Area MultiPark (Multidisciplinary Research in Parkinson’s disease) at Lund University.

This work was supported by resources and effort provided by the Tracy Family SILQ Center (PI: RJB) established by the Tracy Family, Richard Frimel and Gary Werths, GHR Foundation, David Payne, and the Willman Family brought together by The Foundation for Barnes-Jewish Hospital. This work was also supported by resources and effort provided by the Hope Center for Neurological Disorders and the Department of Neurology at the Washington University School of Medicine. This work was also supported by the Clinical, Fluid Biomarker, and Imaging Cores of the Knight ADRC (P30 AG066444 [PI: JCM], P01 AG03991 [PI: JCM], and P01 AG026276 [PI: JCM]) at the Washington University School of Medicine for participant evaluation, samples, and data collection. The mass spectrometry analyses of BioFINDER-2 and Knight ADRC samples were supported by Eisai industry grant to Washington University (PI: KH and RJB) and the Knight ADRC developmental project (PI: NRB) and R01AG070941 (PI: SES).

The funding sources had no role in the design and conduct of the study; in the collection, analysis, interpretation of the data; or in the preparation, review, or approval of the manuscript.

## Online methods

### Participants

#### BioFINDER-2

We assessed 426 participants from the Swedish BioFINDER-2 study (NCT03174938)^19^, with the complete set of CSF biomarkers available. Participants were recruited at Skåne University Hospital and the Hospital of Ängelholm in Sweden. These participants also had amyloid-PET (n=251), tau-PET (n=417), a magnetic resonance imaging (MRI, n=420) and cognitive assessment (n=426). Also, 220 participants had available CSF biomarkers at follow-up (mean time (SD) = 2.05(0.22) years). Inclusion and exclusion criterion for this study has been detailed before^19^. In summary, CU participants do not fulfil criteria for MCI or dementia according to DSM-5^63^. Subjective cognitive decline (SCD) participants were considered as CU, in accordance with the research framework by the National Institute on Aging-Alzheimer’s Association^64^. MCI participants had a MMSE score above 23, they did not fulfil the criteria for major neurocognitive disorder (dementia) according to DSM-5 and performed worse than −1.5 SD in at least one cognitive domain according to age and education stratified test norms. AD dementia was diagnosed according to the DSM-5 criteria for major neurocognitive disorder due to AD and an abnormal biomarker for Aβ pathology was also required. Participants fulfilling the criteria for any other dementia were categorized as non-AD dementias, as previously described.^19^

#### Knight-ADRC

Knight ADRC cohort consisted of community-dwelling volunteers enrolled in studies of memory and aging at Washington University in St. Louis. All Knight ADRC participants underwent a comprehensive clinical assessment that included a detailed interview of a collateral source, a neurological examination of the participant, the Clinical Dementia Rating (CDR^®^)^65^ and the MMSE^66^. Individuals with a CDR of 0.5 or greater were considered to have a dementia syndrome and the probable aetiology of the dementia syndrome was formulated by clinicians based on clinical features in accordance with standard criteria and methods^67^. In Knight-ADRC cohort, participants were categorized as CU if they were scored CDR=0, either Aβ negative or positive (CU− and CU+, respectively); very mild AD patients (CDR=0.5); and mild AD dementia patients (CDR≥1) if clinical syndrome was typical of symptomatic AD. Participants with CDR≥0.5 with different aetiology were assessed as being other dementia patients regardless of their amyloid status.

All participants gave written informed consent and ethical approval was granted by the Regional Ethical Committee in Lund, Sweden and the Washington University Human Research Protection Office, respectively.

### Fluid biomarkers

Measurement of CSF tau species (*i.e.*, pT205/T205, pT217/T217, pT231/T231, MTBR-tau243 and total-tau [the residue 151-153]) was performed at Washington University in both cohorts using the newly developed IP/MS method, as previously detailed^32^. In BioFINDER-2, CSF levels of Aβ42/40 and NfL were measured using the Elecsys platform as previously explained^19^. Aβ positivity was assessed using CSF Aβ42/40 (<0.080), unless otherwise stated, based on Gaussian-mixture modeling. In Knight-ADRC, CSF Aβ42/40 levels were measured as explained previously^28,68^. The CSF Aβ42/40 positivity threshold (0.0673) had the maximum combined sensitivity and specificity in distinguishing amyloid-PET status. CSF NfL was measured with a commercial ELISA kit (UMAN Diagnostics, Umea, Sweden), as explained previously^69^.

### Image acquisition and processing

Image acquisition and processing details from BioFINDER-2 have been previously reported^19^. In brief, amyloid- and tau-PET were acquired using [^18^F]flutemetamol and [^18^F]RO948, respectively. Amyloid-PET binding was measured as SUVR using a neocortical meta-ROI, and with the cerebellar grey as a reference region. Of note, most of the AD dementia patients did not undergo amyloid-PET in BioFINDER-2, due to the study design. For main analyses, tau-PET binding was measured in a temporal meta-ROI^70^, which included, entorhinal, amygdala, parahippocampal, fusiform, inferior temporal, and middle temporal ROIs, using the inferior cerebellar cortex as reference region without partial volume correction. Additionally, tau-PET binding was also measured in regions covering early (Braak I), intermediate (Braak III-IV) and late (Braak V-VI) tau deposition areas^43^. For assessing cortical thickness, T1-weighted anatomical magnetization-prepared rapid gradient echo (MPRAGE) images (1mm isotropic voxels) were used. A cortical thickness meta-ROI was calculated including entorhinal, inferior temporal, middle temporal, and fusiform using FreeSurfer (v.6.0. https://surfer.nmr.mgh.harvard.edu) parcellation, which are areas known to be susceptible to AD-related atrophy^71^.

Methodological details for imaging processing and quantification for the Knight-ADRC cohort have been also previously reported^71,72^. In brief, MPRAGE data were processed using Freesurfer (v.5.3) to generate regions of interest. Amyloid-PET was acquired with either [^11^C]PIB or [^18^F]florbetapir and was quantified in a neocortical meta-ROI using cerebellar grey as a reference region. SUVR values were transformed to Centiloids^73^ to allow direct comparison between tracers using previously validated transformations^74^. [^18^F]flortaucipir ([^18^F]AV1451) was used as a tau-PET tracer and images were quantified in the same temporal meta-ROI as in BioFINDER-2 and assessed as positive if SUVR>1.32 based on previous work. The same additional regions as in BioFINDER-2 were also used to quantify tau-PET binding in early, intermediate, and late tau deposition regions. In all cases, cerebellar grey was used as a reference region. T1-weighted images were used to measure cortical thickness using the same approach as is in the BioFINDER-2 cohort.

### Neuropsychological testing

mPACC and a global cognitive composite were used as the main cognitive outcome in BioFINDER-2 and Knight-ADRC participants, respectively. In BioFINDER-2 participants, the mPACC-5 composite was calculated using mean of z-scores of Alzheimer’s disease assessment scale (ADAS) delayed recall (weighted double), animal fluency, MMSE^66^, and trail making test-A (TMT-A)^75^, as a sensitive measure of early cognitive impairment^76^. Z-scores were calculated with a group of CU− as reference. Further, we also calculated several cognitive composites averaging z-scores of different cognitive tests. For the memory composite we used ADAS delayed and immediate word recall; for the executive function composite, we used TMT-A, TMT-B and symbol digit test; for the language composite we used the animal fluency test and the Boston naming test total score (BNT)^77^; and, finally for the visuo-spatial composite we used the visual object and space perception (VOSP) cube and letters tests.

In Knight-ADRC, the global cognitive composite was created using mean of z-scores of free and cued selective reminding test (FCSRT) free recall^78^, animal fluency, TMT-A and TMT-B. Z-scores were also calculated from a CU− group as a reference. For the executive function composite, we used TMT-A and TMT-B. We could not obtain any other cognitive composite similar to those derived in BioFINDER-2, due to lack of similar tests. However, we selected individual tests to try to recapitulate similar cognitive measures. For memory we used FCSRT, and for language we used animal fluency. No tests were available related visuo-spatial capacity.

### Model creation

Model development was done with SuStaIn^41^ using cross-sectional data of amyloid-positive participants based on CSF Aβ42/40 levels. We selected these participants because we wanted to create a staging model focused on AD. SuStaIn is a machine-learning technique that unravels temporal progression patterns (stages) allowing for multiple different trajectories (subtypes). For our purpose, we used the event-based model^79^ (or mixture SuStaIn^80^), in which the input data is the probability of each biomarker of being abnormal for each participant. In our case, we used a Gaussian-mixture modelling approach (with 2 Gaussians) to obtain these probabilities. With this information, SuStaIn provides the maximum likelihood sequence by which biomarkers become abnormal, and gives a probability for this ordering, for all subtypes. The number of SuStaIn stages is defined by the number of biomarkers provided to the model (*i.e.*, one per biomarker plus a biomarker negative stage). The selection of the optimal number was determined using cross-validation, optimizing on cross-validation based information criterion (CVIC) and out-of-sample log-likelihood was calculated. The optimal number of subtypes was then selected based on these criteria, using the minimal number of subtypes that had the lowest CVIC and higher log-likelihood^41^. In this study, we used pySuStaIn^81^, a Python implementation of the original method (downloaded 08/2022).

In our initial model with BioFINDER-2 participants, we included all biomarkers available and performed the cross-correlation in models with one, two and three subtypes. Although CVIC measures were lower in the three subtypes model, the similar log-likelihood in all three models, supported the one subtype model as the optimal one due to its lower complexity^82^. Upon examining the outcome of this model, we observed that pT217/T217, pT231/T231, and pT181/T181 position certainty was low, as they seemed to compete for the second position (Ext Data Fig. 1A). To avoid stages with low certainty, we decided to try to optimize this model through iterative removal of these biomarkers. All the possible combinations were created (*i.e.*, removing pT217/T217 and/or pT231/T231 and/or pT181/T181) and compared using the CVIC (Ext Data Fig. 1B). We observed that models including only one of these biomarkers (models 5-7) were better than those including two (models 2-4) or all three (model 1). Further, models including pT181/T181 performed worse, and those including pT217/T217 performed better. Thus, the optimal model was selected as that including only pT217/T217 (model 7). Once the biomarkers to be included in the model were selected, we repeated the cross-validation with models up to three subtypes. Comparing CVIC and log-likelihood values, we once again selected the one subtype model as the optimal (Ext Data Fig. 1C). Based on this cross-validated model, we then staged all BioFINDER-2 participants.

For Knight-ADRC, we created the model from the biomarkers selected in the BioFINDER-2 cohort. To investigate whether one subtype was also the selected model, we run SuStaIn and created cross-validated models for one to three subtypes. Again, based on CVIC (subtypes: 1=578.9; 2=598.1; 3=611.0) and log-likelihood (mean: 1=−29.4; 2=−30.4; 3=−31.0), the less complex model (*i.e.*, one subtype) was selected as the optimal. We then staged all Knight-ADRC participants based on this cross-validated model.

As a sensitivity analysis, we compared the levels of the two biomarkers excluded (pT231/T231 and pT181/T181) with those of pT217/T217 in the optimal model. In summary, we observed that these biomarkers followed a similar trajectory across CSF stages as pT217/T217, although with lower increases in the two cohorts (Ext Data Fig. 8A-B), which supports our decision of removing them from the model to have more stable and independent stages.

### Statistical analyses

All biomarkers were z-scored using participants older than 60 from the CU− group as a reference (BioFINDER-2: n=63 and Knight-ADRC: n=71). When necessary, biomarker data was inverted such that higher z-scores related to higher abnormality across all biomarkers. Differences in biomarkers by CSF stages were assessed using Kruskal-Wallis test. Wilcoxon-test was used for *post-hoc* comparisons adjusted for multiple comparisons with false discovery rate (FDR) correction (only differences in consecutive CSF stages are shown in figures). For categorical data (*i.e.*, sex, *APOE* carriership and diagnosis), we used chi-squared tests. LOESS regressions were used to fit the progression of biomarkers abnormalities across the CSF stages. ROC curves were used to assess the utility of CSF stages for predicting amyloid-PET, tau-PET positivity and to compare AD to non-AD objective cognitive impairment (MCI or dementia states). Maximization of Youden’s index was used to select the optimal CSF stage cut-off in each case (*pROC* and *cutpointr* packages were used). For ordinal categories (*i.e.*, A/T status and diagnosis), ordinal logistic regression models were used (*MASS* and *lmr* packages). An equivalent measure to AUC, the c-index, was used to assess the performance of the CSF staging^83^. Confidence intervals were calculated using bootstrapping. Predicted probabilities of the outcome groups per each CSF stages were calculated using the *predict* function, Longitudinal rates of changes were calculated for every participant using linear regression models. One participant with a very negative rate of change in amyloid-PET (z-score< −1.8) was considered an outlier by visual inspection and excluded from the analysis. Kaplan-Meier curves were used to assess clinical progression using *survival* and *survminer* packages. Cox-proportional hazards model were used to calculate the risk of clinical progression adjusting for age and sex in all cases, and further baseline clinical status if necessary.

All analyses were performed with R (v.4.1.0). A two-sided p value < 0.05 was considered statistically significant.

## Extended data

**Ext Data Fig 1:**
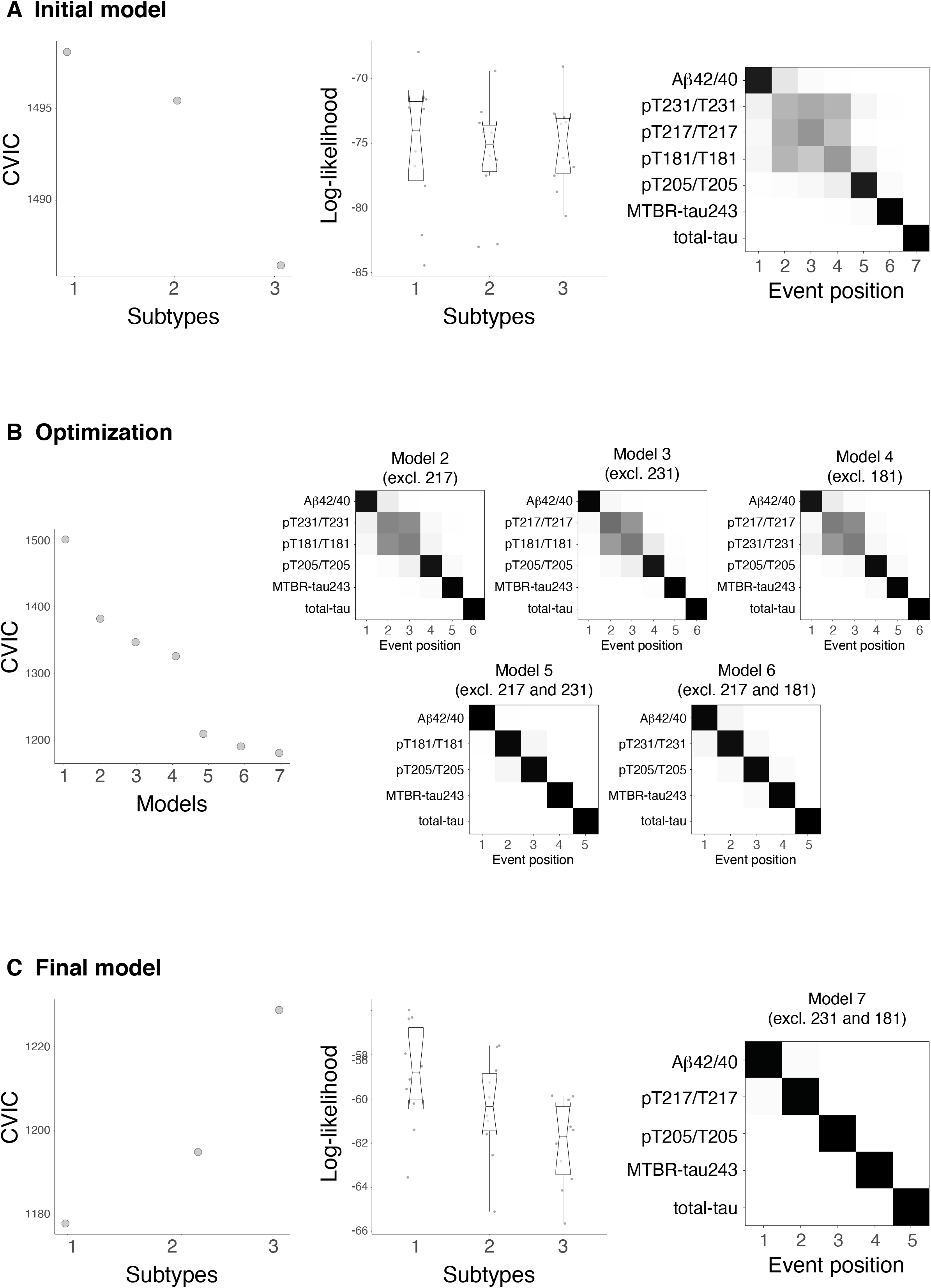
Creation and optimization of the model. Initial model with all CSF biomarkers (Aβ42/40, pT217/T217, pT231/T231, pT181/T181, pT205/T205, MTBR-tau243 and total-tau) is shown in A. First two columns represent the statistics, CVIC and log-likelihood, of this model for one, two and three subtypes. Each dot in log-likelihood plot represents one of the ten cross-validation sets of data. Lower CVIC and higher log-likelihood values represent better performance of the model. Although higher number of subtypes had higher CVIC, the comparable log-likelihood across subtypes suggests that one subtype is complex enough to explain the variability observed in the data. Cross-validated confusion matrix of the one subtype model is shown in the last column. Here, biomarkers are sorted by the time they become abnormal based on the results of SuStaIn. Darkness represents the probability of that biomarker of becoming abnormal at that position, with black being 100%. Given that some biomarkers (pT217/T217, pT231/T231 and pT181/T181) show high overlap on the ordering, we optimized the model by removing these biomarkers systematically (B). All models without one or two of these biomarkers were tested (models 2 to 7). CVIC (left) and cross-validated confusion matrixes (right) for each of these models are shown in B, respectively. CVIC shows that the optimal model was that excluding both pT231/T231 and pT181/T181 (model 7, shown in C). Both CVIC and log-likelihood measures show that one subtype was the optimal model when using this set of biomarkers. Abbreviations: Aβ, amyloid-β; CVIC, cross-validation information criterion; MTBR, microtubule binding region; pT, phosphorylated tau; SuStaIn, subtype and stage inference.

**Ext Data Fig 2:**
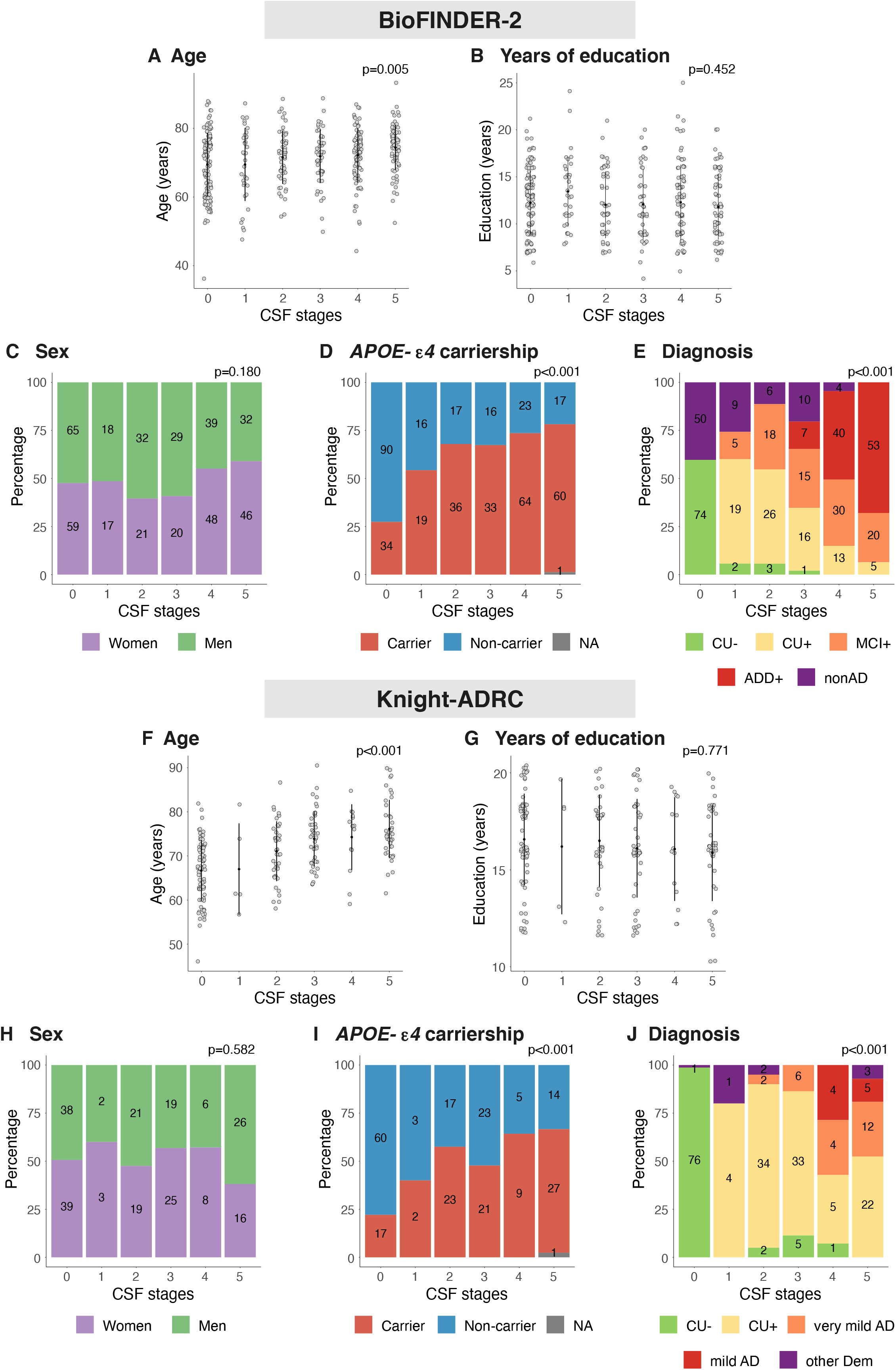
Demographic, genetic and clinical characteristics by CSF stage. Depiction of basic characteristics of BioFINDER-2 (A-E) and Knight-ADRC (F-J) by CSF stage. Kruskal-Wallis or chi-square tests were used to investigate the association between each of these characteristics and CSF stages. P-values of these tests are shown at the top right of each subplot. Number of individuals in each category are shown inside the barplots. Abbreviations: AD, Alzheimer’s disease; ADD+, Alzheimer’s disease dementia amyloid positive; CU−, cognitively unimpaired amyloid negative; CU+, cognitively unimpaired amyloid positive; CSF, cerebrospinal fluid; MCI+, mild cognitive impairment amyloid positive; nonAD, non-Alzheimer’s related disease; other Dem, non-Alzheimer’s type dementia.

**Ext Data Fig 3:**
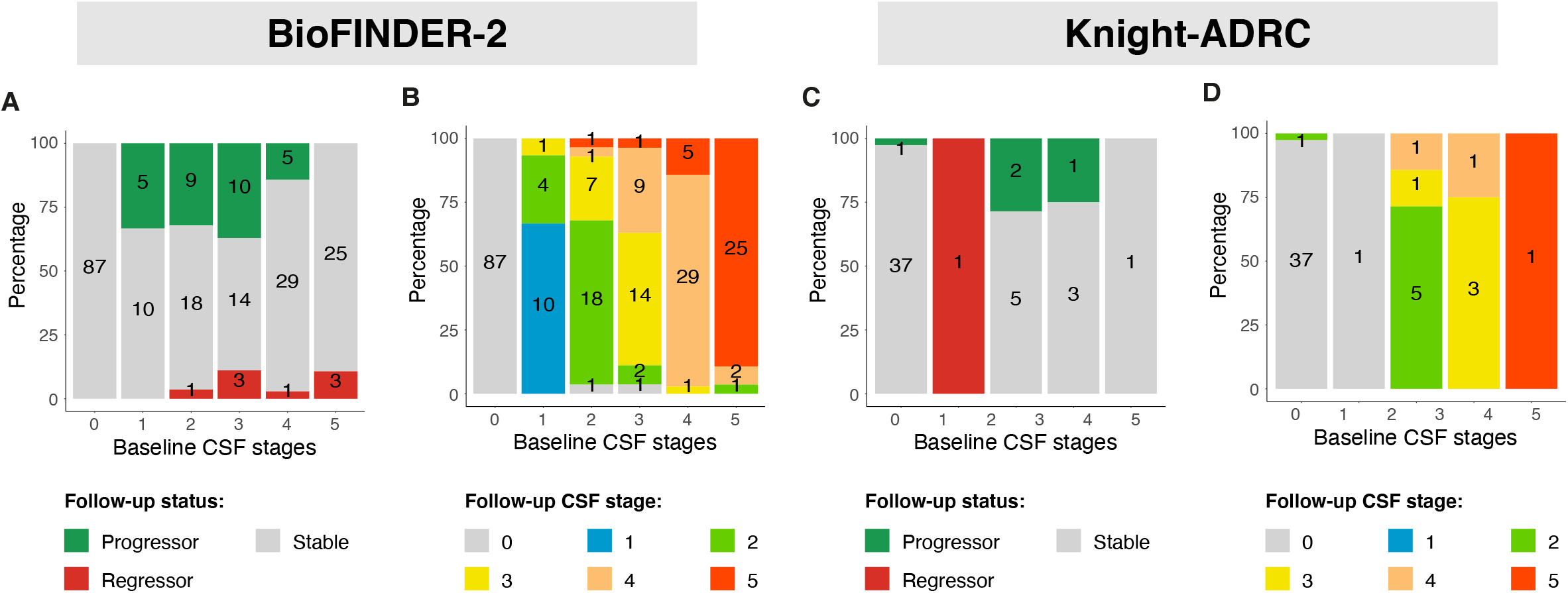
Model stability. Depiction of the evolution of CSF stages in BioFINDER-2 (n=220, A-B) and Knight-ADRC participants (n=51, C-D) with longitudinal CSF available. As longitudinal CSF Aβ42/40 levels were not available for any BioFINDER-2 participant, we imputed this data with their baseline levels. We show the number of progressors, regressors and stable participants in A and C, for each cohort respectively. In B and D, we further show the CSF stages at follow-up. For those Knight-ADRC with more than one longitudinal visit we took the one more distant from the baseline. Abbreviations: Aβ, amyloid-β; CSF, cerebrospinal fluid.

**Ext Data Fig 4:**
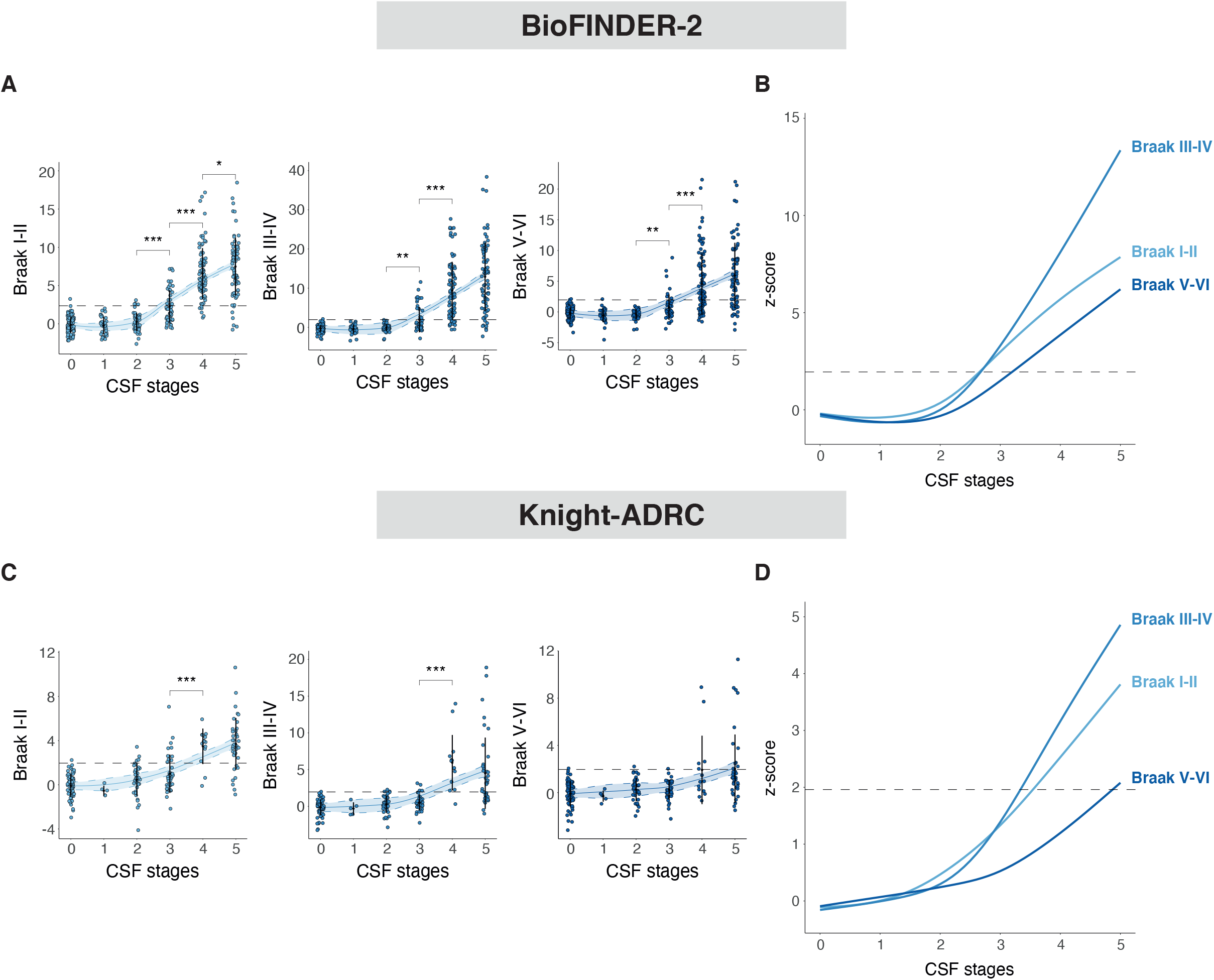
Tau-PET binding in different Braak regions by CSF stages. Depiction of tau-PET binding in different areas of tau deposition, by CSF stage in all BioFINDER-2 (A) and Knight-ADRC participants (B). These areas include regions of early (Braak I-II), intermediate (Braak II-IV) and late (Braak V-VI) tau deposition. Tau-PET levels are z-scored based on a group of CU− participants (BioFINDER-2: n=63 and Knight-ADRC: n=71) and all increases represent increase in abnormality. Significant differences in contiguous CSF stages are shown with asterisks. Horizontal line is drawn at z-score=1.96 which represents 95%CI of the reference group (CU−). Smoothed LOESS lines of all AD biomarkers are shown in B for comparison. CSF stage 0 represent being classified as normal by the model. *: p<0.05; **: p<0.01; ***: p<0.001. Abbreviations: Aβ, amyloid-β; AD, Alzheimer’s disease; CI, confidence interval; CU−, cognitively unimpaired amyloid negative; CSF, cerebrospinal fluid; LOESS, locally estimated scatterplot smoothing; PET, positron emission tomography; ROI, region of interest.

**Ext Data Fig 5:**
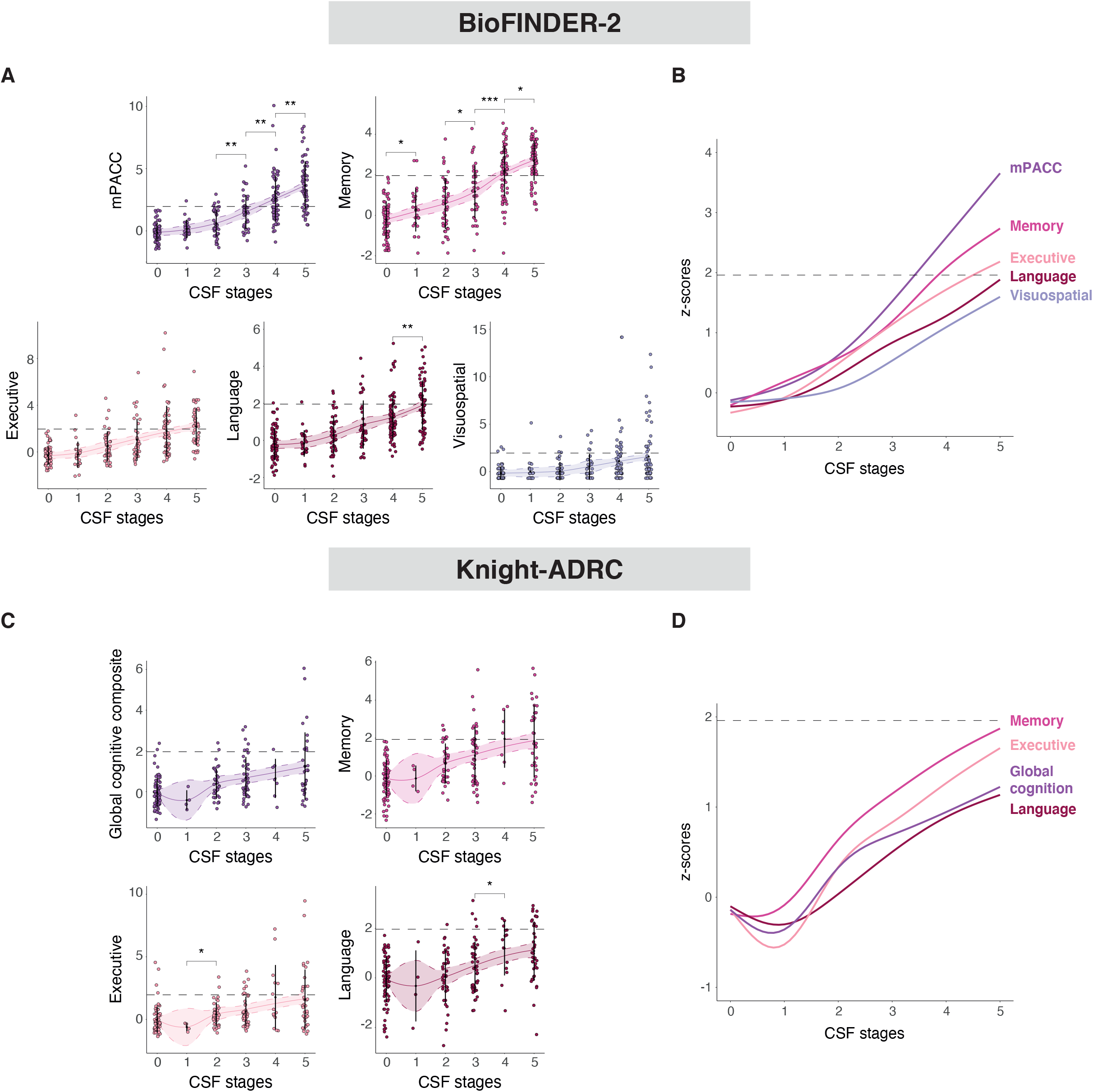
Cognitive composites by CSF stages. Depiction of different cognitive measures, by CSF stage in BioFINDER-2 (A) and Knight-ADRC participants (B). These measures include: mPACC (ADAS-delayed, animal fluency, MMSE and TMT-A), memory (ADAS-delayed and ADAS-immediate), executive function (TMT-A, TMT-B and symbols digit), language (animal fluency and BNT-15) and visuospatial (VOSP-cube and VOSP-incomplete) for BioFINDER-2. For Knight-ADRC we had a global cognitive composite (FCSRT, animals, TMT-A and TMT-B), an executive function composite (TMT-A and TMT-B), a memory (FCSRT) and language (animal fluency) tests. Cognitive scores are z-scored based on a group of CU− participants (BioFINDER-2: n=60 and Knight-ADRC: n=71) and all increases represent increase in abnormality. Significant differences in contiguous CSF stages are shown with asterisks. Horizontal line is drawn at z-score=1.96 which represents 95%CI of the reference group (CU−). Smoothed LOESS lines of all AD biomarkers are shown in B for comparison. We excluded non-AD dementia patients to avoid bias in these analyses. CSF stage 0 represents being classified as normal by the model. *: p<0.05; **: p<0.01; ***: p<0.001. Abbreviations: AD, Alzheimer’s disease; ADAS, Alzheimer’s disease assessment scale; BNT, Boston naming test; CI, confidence interval; CU−, cognitively unimpaired amyloid negative; CSF, cerebrospinal fluid; FCSRT, free and cued selective reminding test; LOESS, locally estimated scatterplot smoothing; MMSE, Mini-Mental state examination; mPACC, modified version of preclinical Alzheimer’s disease cognitive composite; TMT, trial making test; VOSP, visual object and space perception battery.

**Ext Data Fig 6:**
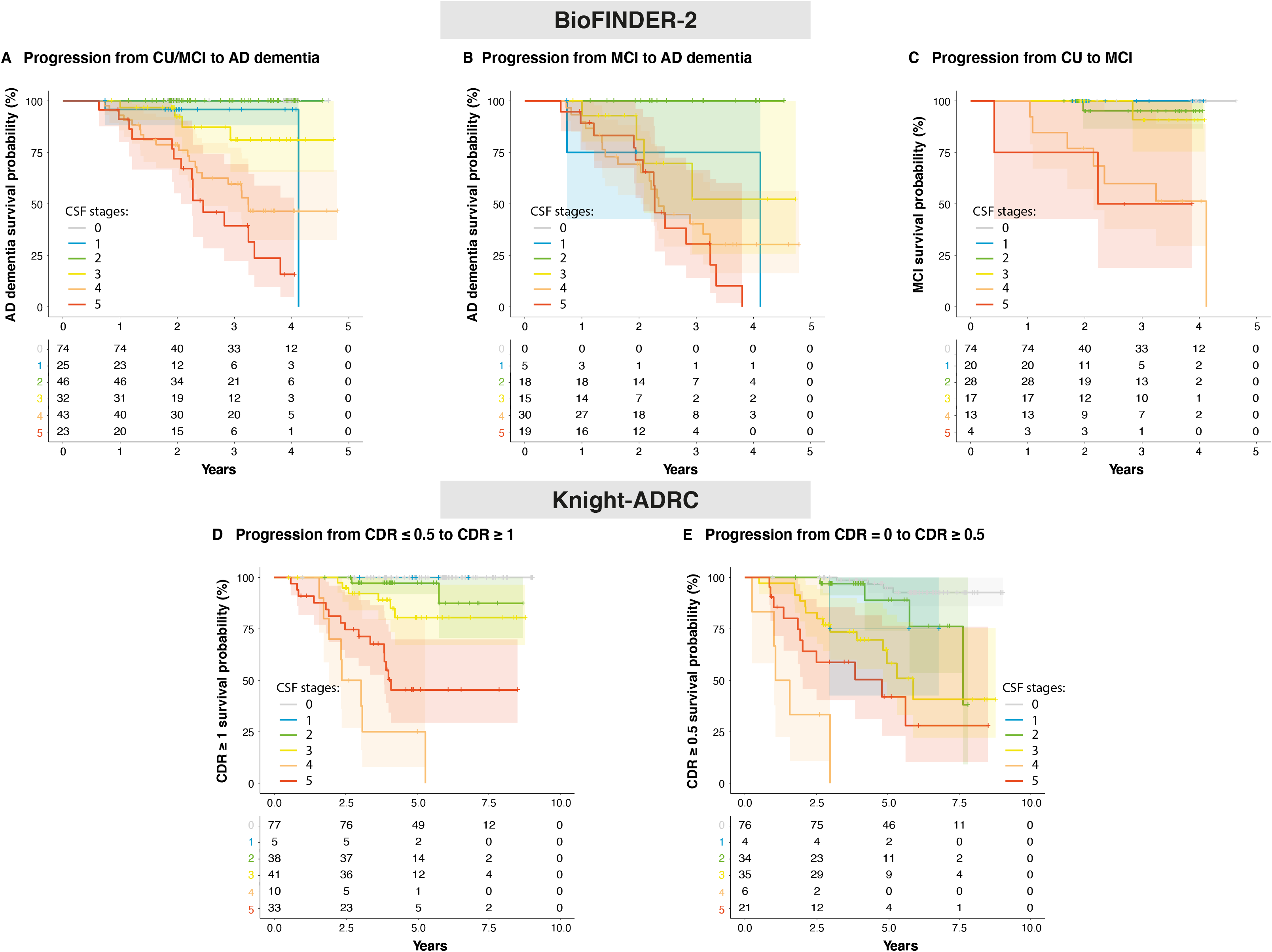
Individual CSF stages for predicting clinical progression. Kaplan-Meier curves for all individual CSF stages in BioFINDER-2 (A-C) and Knight-ADRC (D-E) participants. For BioFINDER-2, progression from CU or MCI at baseline to AD dementia is shown in A; progression from MCI at baseline to AD dementia is shown in B and; progression from CU at baseline to MCI is shown in C. For Knight-ADRC, progression from CDR=0 or CDR=0.5 at baseline to CDR≥1 is shown in D and; progression from CDR=0 at baseline to CDR≥0.5 is shown in E. Abbreviations: AD, Alzheimer’s disease; CDR, clinical dementia rating; CSF, cerebrospinal fluid; CU, cognitively unimpaired; MCI, mild cognitive impairment.

**Ext Data Fig 7:**
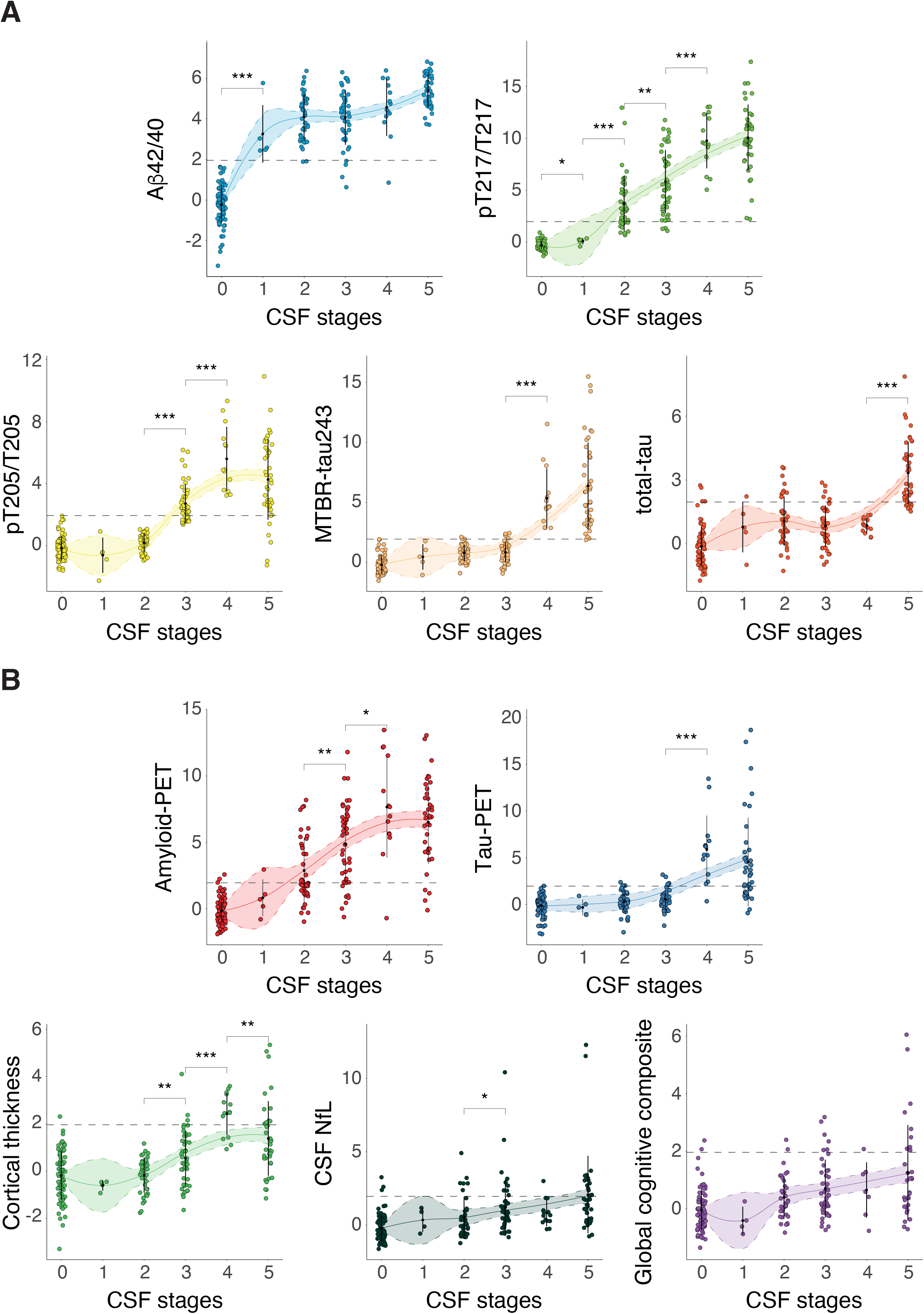
Individual biomarker levels by CSF stage in Knight-ADRC participants. Individual CSF biomarker levels, included in the model, by CSF stage participants are shown in A including all Knight-ADRC participants. Depiction of individual AD-biomarker levels, not used in the creation of the model, per CSF stage are shown in B. All biomarker levels are z-scored based on a group of CU− participants (n=71) and all increases represent increase in abnormality. Significant differences in contiguous CSF stages are shown with asterisks. Horizontal line is drawn at z-score=1.96 which represents 95%CI of the reference group (CU−). CSF stage 0 represent being classified as normal by the model. Black dots and vertical lines represent mean and SD per CSF stage. *: p<0.05; **: p<0.01; ***: p<0.001. Abbreviations: Aβ, amyloid-β; CI, confidence interval; CU−, cognitively unimpaired amyloid negative; CSF, cerebrospinal fluid; MMSE, Mini-Mental state examination; MTBR, microtubule binding region; NfL, neurofilament light; PET, positron emission tomography; pT, phosphorylated tau; SuStaIn, subtype and stage inference.

**Ext Data Fig 8:**
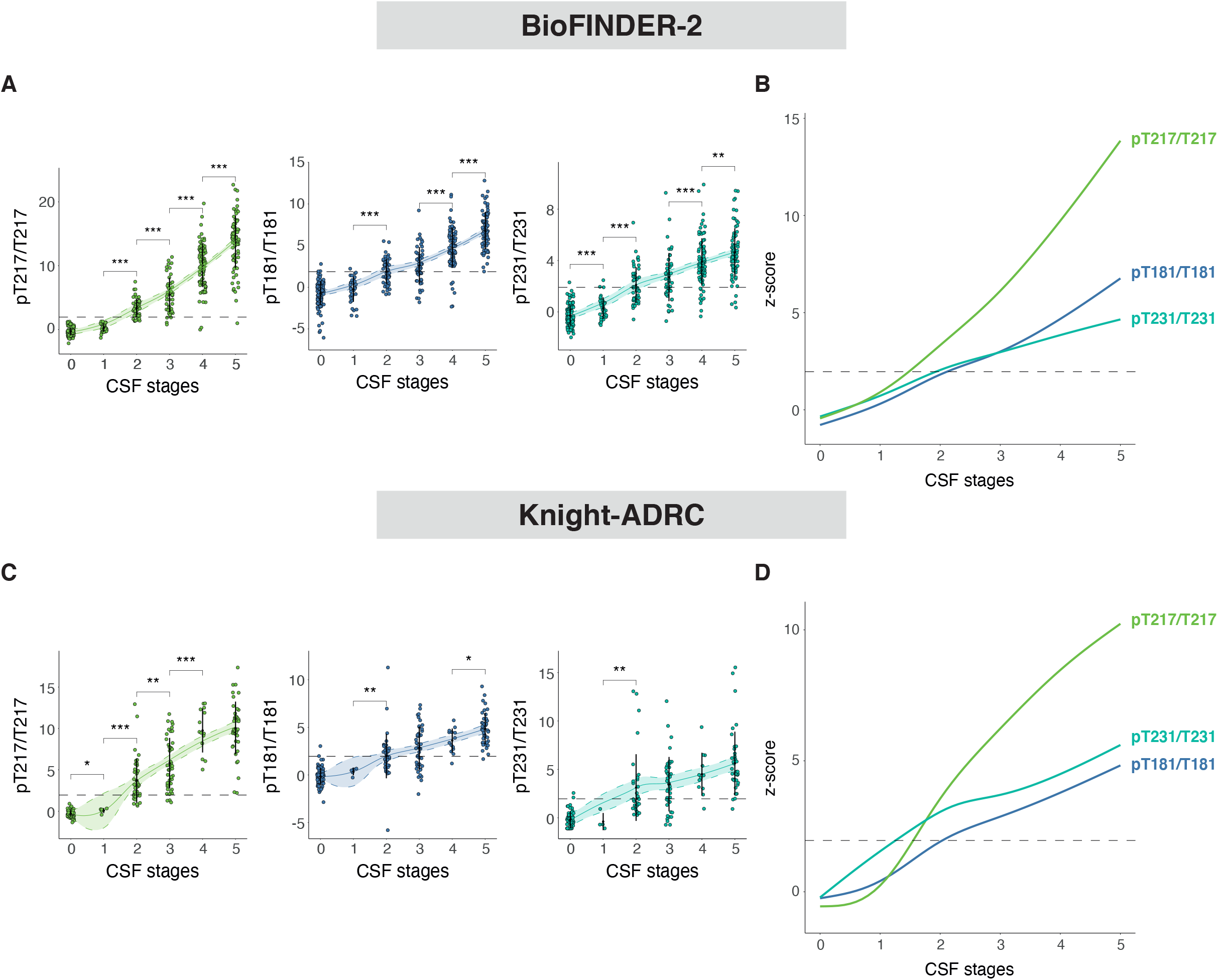
Excluded CSF biomarkers by CSF stage. Depiction of the CSF biomarkers excluded in the optimal model (pT231/T231 and pT181/T181) by CSF stage in BioFINDER-2 (A-B) and Knight-ADRC (C-D) participants. CSF pT217/T217 is also shown for comparison. CSF levels are z-scored based on a group of CU− participants (BioFINDER-2: n=63, Knight-ADRC: n=71) and all increases represent increase in abnormality. Significant differences in contiguous CSF stages are shown with asterisks. Horizontal line is drawn at z-score=1.96 which represents 95%CI of the reference group (CU−). Smoothed LOESS lines of all CSF biomarkers are shown in B (BioFIDNER-2) and D (Knight-ADRC) for comparison. CSF stage 0 represent being classified as normal by the model. Black dots and vertical lines represent mean and SD per CSF stage, respectively. *: p<0.05; **: p<0.01; ***: p<0.001. Abbreviations: Aβ, amyloid-β; CI, confidence interval; CU−, cognitively unimpaired amyloid negative; CSF, cerebrospinal fluid; LOESS, locally estimated scatterplot smoothing; MTBR, microtubule binding region; pT, phosphorylated tau; SuStaIn, subtype and stage inference.

