## Supplementary Material for "Novel CSF tau biomarkers can be used for disease staging of sporadic Alzheimer’s disease"

**Supplementary Data:**

**Supplementary Table 1: Characteristics of BioFINDER-2 participants with follow-up CSF data**

**Supplementary Table 2: Statistics of each CSF biomarker and their differences by CSF stage**

**Supplementary Table 3: Statistics of AD-biomarkers and their differences by CSF stage**

**Supplementary Table 4: Statistics of tau-PET binding in different regions and their differences by CSF stage**

**Supplementary Table 5: Statistics of cognitive composites and their differences by CSF stage**

**Supplementary Table 6: CSF stages for predicting predicting A/T status and as a diagnostic tool**

**Supplementary Table 7: Characteristics of BioFINDER-2 participants with follow-up AD biomarkers**

**Supplementary Table 8: Statistics of AD-biomarkers longitudinal rates of change and their differences by CSF stage**

**Supplementary Table 9: CSF stages for predicting disease progression**

**Supplementary Table 10: Characteristics of Knight-ADRC participants with follow-up CSF data**

| **CSF stage at baseline** | **All (n=220)** | **CU- (n=80)** | **CU+ (n=49)** | **MCI+ (n=46)** | **ADD+ (n=28)** | **nonAD (n=17)** |
| --- | --- | --- | --- | --- | --- | --- |
| **Age, years** | 71.4 (8.80) | 70.7 (9.46) | 72.8 (7.71) | 71.2 (9.06) | 74.1 (7.67) | 67.5 (8.45) |
| **Women, n(%)** | 107 (48.6%) | 39 (48.8%) | 23 (46.9%) | 24 (52.2%) | 15 (53.6%) | 6 (35.3%) |
| ***APOE-ε4* carriershp, n(%)** | 121 (55.0%) | 26 (32.5%) | 36 (73.5%) | 33 (71.7%) | 20 (71.4%) | 6 (35.3%) |
| **Years of education** | 12.2 (3.9) | 12.0 (3.2) | 12.0 (3.5) | 12.9 (5.1) | 11.5 (3.9) | 12.6 (4.5) |
| **Years between baseline and follow-up** | 2.05 (0.22) | 1.91 (0.11) | 2.04 (0.23) | 2.16 (0.24) | 2.19 (0.17) | 2.17 (0.19) |
| **Stage at baseline, n(%)** | | | | | | |
| **0** | 87 (39.5%) | 74 (92.5%) | 0 (0%) | 0 (0%) | 0 (0%) | 13 (76.5%) |
| **1** | 15 (6.8%) | 2 (2.5%) | 10 (20.4%) | 3 (6.5%) | 0 (0%) | 0 (0%) |
| **2** | 28 (12.7%) | 3 (3.8%) | 15 (30.6%) | 9 (19.6%) | 0 (0%) | 1 (5.9%) |
| **3** | 27 (12.3%) | 1 (1.3%) | 12 (24.5%) | 10 (21.7%) | 2 (7.1%) | 2 (11.8%) |
| **4** | 35 (15.9%) | 0 (0%) | 10 (20.4%) | 15 (32.6%) | 9 (32.1%) | 1 (5.9%) |
| **5** | 28 (12.7%) | 0 (0%) | 2 (4.1%) | 9 (19.6%) | 17 (60.7%) | 0 (0%) |
| **Stage at follow-up, n(%)** | | | | | | |
| **0** | 89 (40.5%) | 76 (95.0%) | 0 (0%) | 0 (0%) | 0 (0%) | 13 (76.5%) |
| **1** | 10 (4.5%) | 2 (2.5%) | 5 (10.2%) | 3 (6.5%) | 0 (0%) | 0 (0%) |
| **2** | 25 (11.4%) | 1 (1.3%) | 20 (40.8%) | 3 (6.5%) | 0 (0%) | 1 (5.9%) |
| **3** | 23 (10.5%) | 1 (1.3%) | 9 (18.4%) | 11 (23.9%) | 0 (0%) | 2 (11.8%) |
| **4** | 41 (18.6%) | 0 (0%) | 14 (28.6%) | 15 (32.6%) | 11 (39.3%) | 1 (5.9%) |
| **5** | 32 (14.5%) | 0 (0%) | 1 (2.0%) | 14 (30.4%) | 17 (60.7%) | 0 (0%) |

**Supplementary Table 1: Characteristics of BioFINDER-2 participants with follow-up CSF data**

As CSF Aβ42/40 levels were not available for any participant we imputed this data with their baseline levels. Data is shown as mean (SD) unless otherwise stated.

Abbreviations: Aβ, amyloid-β; AD, Alzheimer’s disease; ADD+, Alzheimer’s disease dementia amyloid positive; CU-, cognitively unimpaired amyloid negative; CU+, cognitively unimpaired amyloid positive; CSF, cerebrospinal fluid; MCI+, mild cognitive impairment amyloid positive; nonAD, non-Alzheimer’s related disease; SD, standard deviation.

| Biomarker | Mean (SD)  CSF stage 0 | Mean (SD)  CSF stage 1 | Mean (SD)  CSF stage 2 | Mean (SD)  CSF stage 3 | Mean (SD)  CSF stage  4 | Mean (SD)  CSF stage  5 |
| --- | --- | --- | --- | --- | --- | --- |
| **BioFINDER-2** | | | | | | |
| Aβ42/40 | 0.02 (0.83)^b,c,d,e,f^ | 2.84 (0.7)^a,c,d,e,f^ | 3.59 (1.06)^a,b,e,f^ | 3.51 (1.42)^a,b,e,f^ | 4.45 (0.94)^a,b,c,d,f^ | 4.91 (0.77)^a,b,c,d,e^ |
| pT217/T217 | -0.3 (0.64)^b,c,d,e,f^ | 0.25 (0.64)^a,c,d,e,f^ | 3.39 (1.38)^a,b,d,e,f^ | 5.47 (2.69)^a,b,c,e,f^ | 10.04 (3.39)^a,b,c,d,f^ | 13.77 (4.09)^a,b,c,d,e^ |
| pT205/T205 | -0.15 (1.13)^c,d,e,f^ | -0.28 (0.95)^c,d,e,f^ | 0.24 (0.99)^a,b,d,e,f^ | 3.64 (1.52)^a,b,c,e,f^ | 5.49 (2.45)^a,b,c,d^ | 5.8 (3.26)^a,b,c,d^ |
| MTBR-tau243 | -0.24 (0.95)^c,d,e,f^ | -0.07 (0.74)^d,e,f^ | 0.31 (1.02)^a,e,f^ | 0.57 (0.92)^a,b,e,f^ | 5.71 (2.58)^a,b,c,d,f^ | 13.33 (7.37)^a,b,c,d,e^ |
| Total-tau | -0.17 (1.05)^b,c,d,e,f^ | 0.38 (1.2)^a,e,f^ | 0.56 (1.19)^a,e,f^ | 0.23 (0.89)^a,e,f^ | 1.54 (0.72)^a,b,c,d,f^ | 5.08 (2.11)^a,b,c,d,e^ |
| **Knight-ADRC** | | | | | | |
| Aβ42/40 | -0.23 (0.94)^b,c,d,e,f^ | 3.26  (1.42)^a,f^ | 4.1  (1.15)^a,f^ | 4.07  (1.34)^a,f^ | 4.57  (1.41)^a^ | 5.39 (0.81)^a,b,c,d^ |
| pT217/T217 | -0.29 (0.38)^b,c,d,e,f^ | 0.08 (0.30)^a,c,d,e,f^ | 3.7 (2.65)^a,b,d,e,f^ | 5.81 (3.05)^a,b,c,e,f^ | 9.76 (2.66)^a,b,c,d^ | 10.05 (3.19)^a,b,c,d^ |
| pT205/T205 | -0.16 (0.78)^c,d,e,f^ | -0.61 (1.14)^d,e,f^ | 0.2 (0.63)^a,d,e,f^ | 2.72 (1.25)^a,b,c,e,f^ | 5.59 (2.06)^a,b,c,d^ | 4.27 (2.59)^a,b,c,d^ |
| MTBR-tau243 | -0.17 (0.74)^c,d,e,f^ | 0.47  (1.06)^e,f^ | 0.80 (0.69)^a,e,f^ | 0.85 (0.85)^a,e,f^ | 5.36 (2.48)^a,b,c,d^ | 6.38 (3.61)^a,b,c,d^ |
| Total-tau | -0.13 (0.92)^c,d,e,f^ | 0.78  (1.19)^f^ | 1.06  (1.16)^a,f^ | 0.83  (0.97)^a,f^ | 0.85  (0.35)^a,f^ | 3.33 (1.48)^a,b,c,d,e^ |

**Supplementary Table 2: Statistics of each CSF biomarker and their differences by CSF stage**

Mean (SD) z-scores by CSF stages are shown in calculated from a group of cognitively unimpaired amyloid negative participants, independently in each cohort. Differences among CSF stages were calculated using pairwise Wilcoxon test. P-values were FDR-corrected for multiple comparisons. Aβ42/40 z-scores were inverted to obtain higher values for higher abnormality. Letters represent statistically significant difference with: a, CSF stage 0; b, CSF stage 1; c, CSF stage 2; d, CSF stage 3; e, CSF stage 4; f, CSF stage 5.

Abbreviations: Aβ, amyloid-β; CSF, cerebrospinal fluid; FDR, false-discovery rate; MTBR, microtubule binding region; pT, phosphorylated tau; SD, standard deviation.

| Biomarker | Mean (SD)  CSF stage 0 | Mean (SD)  CSF stage 1 | Mean (SD)  CSF stage 2 | Mean (SD)  CSF stage 3 | Mean (SD)  CSF stage 4 | Mean (SD)  CSF stage 5 |
| --- | --- | --- | --- | --- | --- | --- |
| **BioFINDER-2** | | | | | | |
| Amyloid-PET | -0.19 (0.58)^b,c,d,e,f^ | 0.93 (2.12)^a,c,d,e,f^ | 4.01 (2.71)^a,b,d,e,f^ | 5.94 (2.64)^a,b,c,e,f^ | 8.3 (3.03)^a,b,c,d^ | 9.06 (3.17)^a,b,c,d^ |
| Tau-PET | -0.27  (0.92)^d,e,f^ | -0.4 (1.12)^d,e,f^ | -0.07 (0.9)^d,e,f^ | 2.02 (3.16)^a,b,c,e,f^ | 9.25 (7.03)^a,b,c,d,f^ | 12.67 (8.43)^a,b,c,d,e^ |
| Cortical thickness | -0.22  (0.99)^d,e,f^ | 0.1 (1.19)^d,e,f^ | 0.08 (1.23)^d,e,f^ | 0.88 (1.54)^a,b,c,f^ | 1.59 (1.69)^a,b,c^ | 2.07 (1.91)^a,b,c,d^ |
| CSF NfL | -0.15  (0.97)^c,d,e,f^ | 0.76 (2.81)^e,f^ | 0.32 (1.64)^a,e,f^ | 0.70 (1.97)^a,e,f^ | 1.06 (2.17)^a,b,c,d,f^ | 1.72 (1.28)^a,b,c,d,e^ |
| mPACC | -0.11  (0.73)^c,d,e,f^ | 0.17 (0.73)^d,e,f^ | 0.61 (1.08)^a,d,e,f^ | 1.53 (1.37)^a,b,c,e,f^ | 2.6 (1.84)^a,b,c,d,f^ | 3.65 (1.92)^a,b,c,d,e^ |
| **Knight-ADRC** | | | | | | |
| Amyloid-PET | -0.15  (0.97)^c,d,e,f^ | 0.84 (1.38)^d,e,f^ | 2.88 (2.47)^a,d,e,f^ | 4.89 (2.95)^a,b,c,e,f^ | 7.66 (3.81)^a,b,c,d^ | 6.52 (3.13)^a,b,c,d^ |
| Tau-PET | -0.14  (1.1)^d,e,f^ | -0.32 (0.84)^e,f^ | 0.3 (1.2)^e,f^ | 0.56 (1.1)^a,e,f^ | 5.89 (3.62)^a,b,c,d^ | 4.54 (4.75)^a,b,c,d^ |
| Cortical thickness | -0.18  (1.02)^d,e,f^ | -0.6  (0.22)^e,f^ | -0.16 (0.75)^d,e,f^ | 0.55 (1.16)^a,c,e,f^ | 2.42 (0.90)^a,b,c,d,f^ | 1.38 (1.58)^a,b,c,d,e^ |
| CSF NfL | -0.22  (0.92)^c,d,e,f^ | 0.35  (0.83) | 0.51 (1.41)^a,d,f^ | 1.19 (1.92)^a,c,f^ | 0.81  (0.90)^a^ | 2.08 (2.65)^a,c,d^ |
| Global cognitive composite | -0.05  (0.71)^c,d,f^ | -0.42  (0.49)^f^ | 0.39  (0.74)^a^ | 0.69  (1.01)^a^ | 0.65  (0.96) | 1.25  (1.66)^a,b^ |

**Supplementary Table 3: Statistics of AD-biomarkers and their differences by CSF stage**

Mean (SD) z-scores by CSF stages are shown calculated from a group of cognitively unimpaired amyloid negative participants, independently in each cohort. Differences among CSF stages were calculated using pairwise Wilcoxon test. P-values were FDR-corrected for multiple comparisons. Cortical thickness and mPACC z-scores were inverted to obtain higher values for higher abnormality. Letters represent statistically significant difference with: a, CSF stage 0; b, CSF stage 1; c, CSF stage 2; d, CSF stage 3; e, CSF stage 4; f, CSF stage 5.

Abbreviations: Aβ, amyloid-β; CSF, cerebrospinal fluid; FDR, false-discovery rate; mPACC, modified preclinical Alzheimer’s cognitive composite; MTBR, microtubule binding region; NfL, neurofilament light; PET, positron emission tomography; pT, phosphorylated tau; SD, standard deviation.

| ROI | Mean (SD)  CSF stage 0 | Mean (SD)  CSF stage 1 | Mean (SD)  CSF stage 2 | Mean (SD)  CSF stage 3 | Mean (SD)  CSF stage 4 | Mean (SD)  CSF stage 5 |
| --- | --- | --- | --- | --- | --- | --- |
| **BioFINDER-2** | | | | | | |
| Braak I-II | -0.14 (1.00)^c,d,e,f^ | -0.23 (1.15)^d,e,f^ | 0.25 (1.18)^a,d,e,f^ | 2.26 (2.14)^a,b,c,e,f^ | 6.58 (3.31)^a,b,c,d,f^ | 7.49 (3.77)^a,b,c,d,e^ |
| Braak III-IV | -0.28 (0.92)^d,e,f^ | -0.41 (1.12)^d,e,f^ | -0.10 (0.9)^d,e,f^ | 1.97 (3.24)^a,b,c,e,f^ | 9.37 (7.35)^a,b,c,d,f^ | 12.92 (8.76)^a,b,c,d,e^ |
| Braak V-VI | -0.19 (0.92)^d,e,f^ | -0.55 (1.13)^d,e,f^ | -0.37 (0.76)^d,e,f^ | 0.77 (2.09)^a,b,c,e,f^ | 4.58 (4.98)^a,b,c,d^ | 5.93 (5.32)^a,b,c,d^ |
| **Knight-ADRC** | | | | | | |
| Braak I-II | -0.07 (1.03)^c,d,e,f^ | -0.54 (0.46)^d,e,f^ | 0.47 (1.47)^a,e,f^ | 1 (1.74)^a,b,e,f^ | 3.48 (1.61)^a,b,c,d^ | 3.70 (2.35)^a,b,c,d^ |
| Braak III-IV | -0.14 (1.10)^d,e,f^ | -0.30 (0.85)^e,f^ | 0.28 (1.17)^e,f^ | 0.53 (1.08)^a,e,f^ | 5.94 (3.78)^a,b,c,d^ | 4.51 (4.85)^a,b,c,d^ |
| Braak V-VI | -0.08 (1.07)^e,f^ | -0.30 (0.47)^f^ | 0.26 (0.99)^f^ | 0.24 (0.85)^f^ | 1.92 (2.9)^a^ | 2.01 (2.91)^a,b,c,d^ |

**Supplementary Table 4: Statistics of tau-PET binding in different regions and their differences by CSF stage**

Mean(SD) z-scores by CSF stages are shown calculated from a group of cognitively unimpaired amyloid negative participants, independently in each cohort. Differences among CSF stages were calculated using pairwise Wilcoxon test. P-values were FDR-corrected for multiple comparisons. Letters represent statistically significant difference with: a, CSF stage 0; b, CSF stage 1; c, CSF stage 2; d, CSF stage 3; e, CSF stage 4; f, CSF stage 5.

Abbreviations: CSF, cerebrospinal fluid; FDR, false-discovery rate; PET, positron emission tomography; ROI, region of interest; SD, standard deviation.

| Biomarker | Mean (SD)  CSF stage 0 | Mean (SD)  CSF stage1 | Mean (SD)  CSF stage 2 | Mean (SD)  CSF stage 3 | Mean (SD)  CSF stage 4 | Mean (SD)  CSF stage 5 |
| --- | --- | --- | --- | --- | --- | --- |
| **BioFINDER-2** | | | | | | |
| mPACC | -0.11 (0.73)^c,d,e,f^ | 0.17  (0.73)^d,e,f^ | 0.61  (1.08)^a,d,e,f^ | 1.53 (1.37)^a,b,c,e,f^ | 2.6 (1.84)^a,b,c,d,f^ | 3.65 (1.92)^a,b,c,d,e^ |
| Memory | -0.22 (0.89)^b,c,d,e,f^ | 0.27  (1.07)^a,d,e,f^ | 0.58  (1.25)^a,d,e,f^ | 1.17 (1.31)^a,b,c,e,f^ | 2.18 (1.24)^a,b,c,d,f^ | 2.7 (0.98)^a,b,c,d,e^ |
| Executive function | -0.29 (0.78)^c,d,e,f^ | -0.24  (1.13)^c,d,e,f^ | 0.49  (1.27)^a,b,e,f^ | 1.04 (1.8)^a,b,f^ | 1.79 (2.16)^a,b,c^ | 2.15 (1.58)^a,b,c,d^ |
| Language | -0.20  (0.86)^c,d,e,f^ | -0.17  (0.71)^d,e,f^ | 0.28  (1.02)^a,e,f^ | 0.85 (1.3)^a,b,f^ | 1.22 (1.18)^a,b,c,f^ | 1.91 (1.32)^a,b,c,d,e^ |
| Visuospatial | -0.15  (0.69)^d,e,f^ | 0.00  (1.17)^e,f^ | 0.06  (0.96)^e,f^ | 0.54 (1.34)^a^ | 1.11 (2.63)^a,b,c^ | 1.59 (2.82)^a,b,c^ |
| MMSE | -0.14 (0.91)^c,d,e,f^ | 0.19  (1.26)^d,e,f^ | 0.68  (1.48)^a,d,e,f^ | 2.17 (2.8)^a,b,c,e,f^ | 4.2 (3.98)^a,b,c,d,f^ | 6.15 (4.51)^a,b,c,d,e^ |
| **Knight-ADRC** | | | | | | |
| Global cognitive composite | -0.05  (0.71)^c,d,f^ | -0.42  (0.49)^f^ | 0.39  (0.74)^a^ | 0.69 (1.01)^a^ | 0.65 (0.96) | 1.25 (1.66)^a,b^ |
| Memory | -0.12 (0.98)^c,d,e,f^ | -0.13  (0.68) | 0.68  (1.04)^a,f^ | 1.07 (1.53)^a^ | 1.96 (1.53)^a^ | 1.84 (1.92)^a,c^ |
| Executive function | -0.01 (1.03)^c,d,e,f^ | -0.61  (0.33)^c,d,f^ | 0.42  (0.95)^a,b^ | 0.64 (1.18)^a,b^ | 1.77 (2.58)^a^ | 1.59 (2.4)^a,b^ |
| Language | -0.05 (0.92)^e,f^ | -0.32  (1.43) | 0.05  (1.08)^e,f^ | 0.43 (1.03)^e,f^ | 1.2 (1.1)^a,c,d^ | 1.09 (1.19)^a,c,d^ |
| MMSE | -0.02 (1.02)^e,f^ | -0.69  (0.00)^e,f^ | 0.42  (1.27)^e,f^ | 0.4 (1.8)^e,f^ | 3.83 (4.37)^a,b,c,d^ | 3.1 (3.89)^a,b,c,d^ |

**Supplementary Table 5: Statistics of cognitive composites and their differences by CSF stage**

Mean(SD) z-scores by CSF stages are shown calculated from a group of cognitively unimpaired amyloid negative participants, independently in each cohort. Differences among CSF stages were calculated using pairwise Wilcoxon test. P-values were FDR-corrected for multiple comparisons. For all tests higher z-scores represent higher abnormality. Letters represent statistically significant difference with: a, CSF stage 0; b, CSF stage 1; c, CSF stage 2; d, CSF stage 3; e, CSF stage 4; f, CSF stage 5.

Abbreviations: Aβ, amyloid-β; CSF, cerebrospinal fluid; FDR, false-discovery rate; MMSE, Mini-Mental state examination; mPACC, modified preclinical Alzheimer’s cognitive composite; MTBR, microtubule binding region; NfL, neurofilament light; PET, positron emission tomography; pT, phosphorylated tau; SD, standard deviation.

|  | **AUC [95%CI]**  **or**  **C-index[95%CI]** | **CSF stage cut-off (≥)** | **Sensitivity** | **Specificity** | **Accuracy** |
| --- | --- | --- | --- | --- | --- |
| **BioFINDER-2** | | | | | |
| Amyloid-PET | 0.96[0.93,0.98] | 2 | 0.93 | 0.89 | 0.92 |
| Tau-PET | 0.95[0.93,0.97] | 4 | 0.91 | 0.92 | 0.91 |
| A/T status | 0.95[0.93,0.97] | - | - | - | - |
| Diagnosis  (AD *continuum*) | 0.88[0.86,0.91] | - | - | - | - |
| AD vs non-AD cognitive impairment | 0.95[0.93,0.98] | 2 | 0.97 | 0.75 | 0.91 |
| **Knight-ADRC** | | | | | |
| Amyloid-PET | 0.89[0.85,0.94] | 2 | 0.91 | 0.81 | 0.87 |
| Tau-PET | 0.94[0.91,0.96] | 4 | 0.92 | 0.87 | 0.88 |
| A/T status | 0.89[0.86,0.92] | - | - | - | - |

**Supplementary Table 6: CSF stages for predicting predicting A/T status and as a diagnostic tool**

ROC curves were used to characterize CSF staging for predicting amyloid-PET (A), tau-PET (T) positivity or as a diagnostic tool (AD vs non-AD cognitive impairment). Maximization of Youden’s index was used to find the optimal CSF stage for separating groups. In AD vs non-AD analysis, only impaired participants were included (MCI or dementia). Ordinal logistic regression was used to classify A/T status and diagnosis. A-T+ (n=1 in each cohort) and non-AD participants were excluded from A/T status and diagnosis analyses, respectively. A/T status was categorized based on PET. For Knight-ADRC, CSF stages 0 and 1 were merged for the A/T status analysis due to low number of subjects at CSF stage 1. AUC was used in ROC analyses and C-index was used in ordinal logistic regression as a performance measure. Amyloid-PET was considered positive if SUVR>1.03 (BioFINDER-2) and Centiloid>20 (Knight-ADRC), tau-PET was considered positive if SUVR at meta-temporal ROI (Braak I-IV) was higher than 1.36 (BioFINDER-2) and 1.32 SUVR (Knight-ADRC).

Abbreviations: AD, Alzheimer’s disease; A-T+, amyloid-negative tau-positive; AUC, area under the curve; CI, confidence interval; C-index, concordance index; CDR, clinical dementia rating; CSF, cerebrospinal fluid; MCI, mild cognitive impairment; PET, positron emission tomography; ROC, receiver operating characteristic; ROI, region of interest; SUVR, standardized uptake value ratio.

|  | **All (n=399)** | **CU-**  **(n=80)** | **CU+ (n=76)** | **MCI+ (n=86)** | **ADD+ (n=90)** | **non-AD (n=67)** |
| --- | --- | --- | --- | --- | --- | --- |
| **Age, years** | 71.4 (8.45) | 70.7 (9.46) | 71.1 (9.30) | 71.9 (7.48) | 72.6 (6.88) | 70.3 (9.24) |
| **Women, n(%)** | 196 (49.1%) | 39 (48.8%) | 37 (48.7%) | 37 (43.0%) | 50 (55.6%) | 33 (49.3%) |
| ***APOE-ε4* carriership, n(%)^a^** | 228 (57.1%) | 26 (32.5%) | 56 (73.7%) | 60 (69.8%) | 65 (72.2%) | 21 (31.3%) |
| **Years of education^b^** | 12.3 (3.81) | 12.0 (3.22) | 12.2 (3.40) | 12.7 (4.53) | 12.0 (3.98) | 12.8 (3.66) |
| **Amyloid-PET rate^c^** | 0.27 (0.39) | 0.06 (0.19) | 0.43 (0.26) | 0.36 (0.51) | - | 0.20 (0.55) |
| **Follow-up time amyloid-PET, years** | 2.63 (1.01) | 2.81 (1.05) | 2.72 (0.98) | 2.38 (0.96) | - | 2.44 (1.13) |
| **Number follow-ups, amyloid-PET [range]** | 2.39 (0.50) [2-4] | 2.46 (0.50) [2-3] | 2.44 (0.53) [2-4] | 2.29 (0.46) [2-3] | - | 2.33 (0.58) [2-3] |
| **Tau-PET rate^d^** | 0.55 (0.98) | 0.04 (0.22) | 0.27 (0.43) | 0.93 (1.19) | 1.45 (1.30) | 0.13 (0.35) |
| **Follow-up time tau-PET, years** | 2.46 (0.97) | 2.80 (1.07) | 2.88 (1.02) | 2.40 (0.91) | 1.73 (0.30) | 2.07 (0.53) |
| **Number follow-ups, tau-PET [range]** | 2.50 (0.68) [2-4] | 2.59 (0.67) [2-4] | 2.70 (0.71) [2-4] | 2.57 (0.79) [2-4] | 2.23 (0.43) [2-3] | 2.18 (0.39) [2-3] |
| **Cortical thickness rate^e^** | 0.258 (0.374) | 0.041 (0.167) | 0.101 (0.195) | 0.293 (0.285) | 0.716 (0.360) | 0.248 (0.501) |
| **Follow-up time cortical thickness, years** | 2.43 (0.93) | 2.77 (1.03) | 2.85 (0.98) | 2.40 (0.89) | 1.72 (0.31) | 2.03 (0.44) |
| **Number follow-ups, cortical thickness [range]** | 2.51 (0.66) [2-4] | 2.61 (0.64) [2-4] | 2.71 (0.72) [2-4] | 2.60 (0.76) [2-4] | 2.26 (0.45) [2-3] | 2.14 (0.36) [2-3] |
| **mPACC rate^f^** | -0.39 (0.79) | 0.05 (0.27) | 0.07 (0.41) | 0.41 (0.58) | 1.22 (1.21) | 0.51 (0.87) |
| **Follow-up time mPACC, years** | 2.55 (0.94) | 2.78 (1.01) | 2.99 (0.87) | 3.00 (0.84) | 2.03 (0.58) | 1.91 (0.70) |
| **Number follow-ups, mPACC[range]** | 2.95 (0.85) [2-5] | 2.43 (0.50) [2-3] | 3.20 (1.08) [2-5] | 3.65 (0.84) [2-5] | 2.78 (0.42) [2-5] | 2.64 (0.60) [2-5] |

**Supplementary Table 7: Characteristics of BioFINDER-2 participants with follow-up AD biomarkers**

Data is shown as mean (SD) unless otherwise stated. All rates of change are given as z-scores and in all cases higher values represent higher abnormality.

^a^, 1 participant missing; ^b^, 4 participants missing; ^c^, 181 participants missing; ^d^, 87 participants missing; ^e^, 99 participants missing; ^f^, 57 participants missing.

Abbreviations: Aβ, amyloid-β; AD, Alzheimer’s disease; ADD+, Alzheimer’s disease dementia amyloid positive; CU-, cognitively unimpaired amyloid negative; CU+, cognitively unimpaired amyloid positive; CSF, cerebrospinal fluid; MCI+, mild cognitive impairment amyloid positive; mPACC, modified preclinical Alzheimer’s cognitive composite; nonAD, non-Alzheimer’s related disease; PET, positron emission tomography; SD, standard deviation; SUVR, standardized uptake value ratio.

| Biomarker | Mean (SD)  CSF stage 0 | Mean (SD)  CSF stage 1 | Mean (SD)  CSF stage 2 | Mean (SD)  CSF stage 3 | Mean (SD)  CSF stage 4 | Mean (SD)  CSF stage 5 |
| --- | --- | --- | --- | --- | --- | --- |
| Amyloid-PET | 0.03 (0.17)^b,c,d,e^ | 0.36  (0.30)^a,c^ | 0.56 (0.25)^a,b,e,f^ | 0.44  (0.27)^a^ | 0.36  (0.33)^a,c^ | 0.23  (0.54)^c^ |
| Tau-PET | 0.05 (0.24)^d,e,f^ | 0.15 (0.27)^d,e,f^ | 0.18 (0.46)^d,e,f^ | 0.46 (0.75)^a,b,c,e,f^ | 1.38 (1.25)^a,b,c,d^ | 1.18 (1.25)^a,b,c,d^ |
| Cortical thickness | 0.09 (0.29)^d,e,f^ | 0.13 (0.38)^d,e,f^ | 0.07 (0.18)^d,e,f^ | 0.30 (0.35)^a,b,c,e,f^ | 0.44 (0.39)^a,b,c,d^ | 0.55 (0.35)^a,b,c,d^ |
| mPACC | 0.16 (0.50)^d,e,f^ | 0.05 (0.41)^d,e,f^ | 0.13 (0.66)^d,e,f^ | 0.40 (0.66)^a,b,c,e,f^ | 0.80 (0.91)^a,b,c,d^ | 0.83 (1.12)^a,b,c,d^ |

**Supplementary Table 8: Statistics of AD-biomarkers longitudinal rates of change and their differences by CSF stage**

Mean(SD) z-scores by CSF stages are shown in calculated from a group of cognitively unimpaired amyloid negative participants. Differences among CSF stages were calculated using pairwise Wilcoxon test. mPACC and cortical thickness z-scores were inverted to obtain higher values for higher abnormality. P-values were FDR-corrected for multiple comparisons.

Abbreviations: Aβ, amyloid-β; CSF, cerebrospinal fluid; FDR, false-discovery rate; mPACC, modified preclinical Alzheimer’s cognitive composite; PET, positron emission tomography; SD, standard deviation.

|  | **HR [95%CI]** | **p** |
| --- | --- | --- |
| **BioFINDER-2** | | |
| CU & MCI to AD dementia | 5.8 [2.4 – 14.3] | <0.001 |
| MCI to AD dementia | 4.5 [1.8 – 10.8] | <0.001 |
| CU to MCI | 33.1 [6.5 - 169.0] | <0.001 |
| **Knight-ADRC** | | |
| CDR=0 & CDR=0.5 to CDR≥1 | 10.4 [4.2 – 25.6] | <0.001 |
| CDR=0 to CDR≥0.5 | 6.4 [3.1 – 13.2] | <0.001 |

**Supplementary Table 9: CSF stages for predicting disease progression**

Hazard ratios for predicting disease progression for higher CSF stages (4-5) compared to lower positive CSF stages (1-3). Models were adjusted for age and sex in all cases, and additionally disease status at baseline (*i.e.*, CU/MCI in BioFINDER-2 or CDR=0/0.5 in Knight-ADRC), if appropriate.

Abbreviations: AD, Alzheimer’s disease; CDR, clinical dementia rating; CU, cognitively unimpaired; CSF, cerebrospinal fluid; MCI, mild cognitive impairment.

| **CSF stage at baseline** | **All (n=51)** | **CU-**  **(n=37)** | **CU+**  **(n=11)** | **Other dementias (n=3)** |
| --- | --- | --- | --- | --- |
| **Age, years** | 69.0 (6.5) | 68.1 (6.4) | 71.5 (6.9) | 70.1 (6.0) |
| **Women, n(%)** | 25 (49.0%) | 17 (45.9%) | 6 (54.5%) | 2 (66.7%) |
| ***APOE-ε4* carriershp, n(%)** | 18 (35.3%) | 9 (24.3%) | 8 (72.7%) | 1 (33.3%) |
| **Years of education** | 16.2 (2.32) | 16.0 (2.48) | 16.5 (1.51) | 17.3 (3.06) |
| **Years between baseline and follow-up** | 2.84 (0.73) | 2.91 (0.72) | 2.62 (0.86) | 2.88 (0.12) |
| **CSF stage at baseline, n(%)** | | | | |
| **0** | 38 (74.5%) | 36 (97.3%) | 0 (0%) | 2 (66.7%) |
| **1** | 1 (2.0%) | 0 (0%) | 1 (9.1%) | 0 (0%) |
| **2** | 7 (13.7%) | 1 (2.7%) | 5 (45.5%) | 1 (33.3%) |
| **3** | 4 (7.8%) | 0 (0%) | 4 (36.4%) | 0 (0%) |
| **4** | 0 (0%) | 0 (0%) | 0 (0%) | 0 (0%) |
| **5** | 1 (2.0%) | 0 (0%) | 1 (9.1%) | 0 (0%) |
| **CSF stage at follow-up, n(%)** | | | | |
| **0** | 38 (74.5%) | 37 (97.4%) | 1 (100%) | 0 (0%) |
| **1** | 0 (0%) | 0 (0%) | 0 (0%) | 0 (0%) |
| **2** | 6 (11.8%) | 1 (2.6%) | 0 (0%) | 5 (71.4%) |
| **3** | 4 (7.8%) | 0 (0%) | 0 (0%) | 1 (14.3%) |
| **4** | 2 (3.9%) | 0 (0%) | 0 (0%) | 1 (14.3%) |
| **5** | 1 (2.0%) | 0 (0%) | 0 (0%) | 0 (0%) |

**Supplementary Table 10: Characteristics of Knight-ADRC participants with follow-up CSF data**

Data is shown as mean (SD) unless otherwise stated. For those Knight-ADRC with more than one longitudinal visit we took the one more distant from the baseline.

Abbreviations: CU-, cognitively unimpaired amyloid negative; CSF, cerebrospinal fluid; MCI+, mild cognitive impairment amyloid positive; SD, standard deviation.
